# Spatial and temporal associations between animal ownership and malaria prevalence in Africa using cross-sectional national Demographic and Health Surveys

**DOI:** 10.64898/2026.06.05.26355017

**Authors:** Hillary M. Topazian, Camille E. Morgan, Varun Goel

## Abstract

Use of zooprophylaxis as a malaria control strategy has been recommended historically, but a complex relationship exists between animal ownership and malaria infection, with mixed associations described in the literature. We sought to characterize this relationship spatially and temporally in malaria-endemic regions of Africa. We used data from 392,843 individuals from 66 Demographic and Health surveys from countries within Africa to investigate the association between household animal ownership and *Plasmodium* infection. We used Bayesian models with Integrated Nested Laplace Approximation to incorporate spatially varying coefficient processes, allowing the association of interest to vary over space, time, and within strata of vector species occurrence, land cover, and number of animals owned by households. Spatially varying intercept models showed that ownership of cattle, chickens/poultry, goats, horses/donkeys/mules, pigs, and sheep was broadly associated with malaria infection, with odds ratios ranging from 1.55 to 1.67. However, spatially varying slope models revealed considerable heterogeneity, with odds ratio estimates for all animal types demonstrating both protective and harmful effects varying from 0.33 to 3.33 both subnationally and across time. We found no evidence that modification by vector species, number of animals owned, and land cover fully explained the variation in estimates. Unobserved localized cultural, behavioral, or ecological factors likely modify the association between animal ownership and malaria prevalence. Further exploring the nature of this relationship over space and time will be important to understanding how context-specific One Health dynamics between humans, animals and the environment affect malaria prevention and control efforts.

## INTRODUCTION

As of 2023, 94% of the globe’s 263 million malaria cases and 95% of the 597,000 malaria deaths occurred in the World Health Organization’s (WHO) Africa region [1]. Many behavioral and environmental risk factors for malaria have been identified, including rurality, poverty, lack of bed net use, low education, deforestation, and pooling water [2,3]. Research to date has considered transmission at the human-human or human-environmental interface. However, apart from the zoonotic malaria parasite *Plasmodium knowlesi* [4], less is established about the contribution of human-animal interactions, the third critical component of a holistic One Health approach.

Human malaria parasites are transmitted through the bite of the female *Anopheles* mosquito, and thrive in hundreds of *Anopheles* species, 41 of which are considered dominant vectors across different geographies [5]. *Anopheles* differ widely in their preferences to bite indoors (endophagic) vs. outdoors (exophagic), rest indoors (endophilic) vs. outdoors (exophilic), bite during the day or at night, and preference to feed on animals (zoophilic) vs. humans (anthropophilic) [5]. Africa hosts seven dominant vectors species, with *An. arabiensis, An. funestus*, and *An. gambiae* covering wide swaths of the continent [5]. All three are considered anthropophilic, with *An. arabiensis* also considered zoophilic. All three species are also considered exophagic, with *An. funestus* and *An. gambiae* also exhibiting endophagic behavior [5]. However, feeding preferences have been observed to vary across Africa, even within vector species.

*Anopheles* mosquitoes feed on a variety of animals in addition to humans, including birds, cattle, dogs, horses, pigs, and sheep [6]. Household ownership of animals has a varied global distribution, and livestock ownership is widespread across Africa [7]. In Africa, livestock are important to both country economies and individual livelihoods. Livestock make up 27% of agricultural net output and 90% of agricultural production comes from small farms [8]. In half of countries surveyed over the last two decades, over 50% of households owned farm animals [9].

Given the role of livestock among human populations in Africa, zooprophylaxis, described as the use of livestock to divert biting mosquitoes away from humans and towards dead-end animal hosts, is one potential strategy to prevent continuation of the malaria parasite lifecycle [10]. Early WHO passive zooprophylaxis recommendations included strategically placing animals between mosquito breeding sites and human settlements in areas with zoophilic vector species [11]. Forms of active zooprophylaxis include ivermectin-treated cattle [12] and insecticide-treated livestock [13].

A complex relationship exists between livestock ownership and risk of malaria, with positive [14], negative [15], and null [16] associations described in the literature. These relationships vary by country and livestock type, including evidence for zoopotentiation [10], where livestock are associated with increased malaria transmission. However, most of these studies are confined to small geographic areas or a single point in time, making it difficult to study heterogeneity in vector populations, geographic space, and to assess if estimates hold across time. Mathematical modeling supports both zooprophylaxis and zoopotentiation hypotheses: increasing the number of animals in an area can divert biting mosquitoes away from humans, but animals can also amplify the number of parasites by increasing vector survival through a greater availability of blood meals [17]. More analysis of field data is needed to identify areas which might be amenable to zooprophylactic interventions, and the contexts in which those interventions may work best.

In this study, we use cross-sectional multi-country, multi-year data across Africa from the Demographic and Health Surveys (DHS) Program and Bayesian hierarchical models to assess the relationship between malaria infection and household ownership of six common types of livestock. Furthermore, we examine how malaria and household livestock ownership associations vary across space and time and what factors may modify those associations. Use of these methods allows for study of this One Health topic on a large geographic and temporal scale and can inform the identification of areas which may be suitable for further analysis of zooprophylactic strategies.

## METHODS AND METHODS

### Study design and population

We used population-based, nationally representative, cross-sectional studies conducted by the DHS Program to investigate the association between animal ownership and *Plasmodium* infection [18]. Standard DHS surveys collect data on relative wealth, housing, sanitation, education, HIV, maternal health, nutrition, and for malaria, use of insecticide-treated nets, fever, and use of other interventions. Malaria Indicator Surveys (MIS) collect data on a smaller set of health characteristics unique to malaria, conduct malaria testing among children to estimate national prevalence rates, and are often timed around high transmission seasons. AIDS indicator surveys (AIS) collect data on indicators for monitoring national HIV/AIDS programs. We retrieved all available surveys for Africa that tested individuals for malaria with rapid diagnostic tests (RDTs). Individual surveys are referred to by their survey codes which consist of a country code, year, and survey type.

The study outcome was an RDT-based diagnosis of malaria infection, classified as a binary variable. The exposure was household ownership of various animals. The DHS standard survey template generally includes counts for the number of cattle, cows/bulls, chickens/poultry, goats, horses/donkeys/mules, and sheep owned by a household. Animal count variables were re-classified as binary variables for the primary analysis. Further information on survey selection and variable classification can be found in the Supplemental Technical Methods.

### Covariate selection

Individual-level risk factors were selected as covariates using directed acyclic graphs (**Figure S4**) and known relationships between malaria and livestock ownership. These included gender (female/male), residence (rural/urban), household wealth index (five quintiles, modelled continuously), and whether the individual slept under a long-lasting insecticide treated net (“bed net”) the night before the survey [10,19,20]. Gender and bed net use were self-reported at the time of survey administration, residence (urban/rural) was classified at the cluster level by country administrators, and wealth index was categorized by the DHS into quintiles based on a principle components analysis of household attributes [21]. The 2009 Uganda MIS (UG2009MIS) was missing data on bed net use, and data on if the individual’s household has at least one mosquito net was used as a proxy variable instead.

Informed consent was obtained by the DHS program from all participants prior to data collection in accordance with the ethical guidelines from the DHS program and with approval from the Institutional Review Boards of ICF and participating countries [22].

### Spatial, temporal, and ecological variables

GPS coordinates for the center of each survey cluster are collected by the DHS program and offset by up to 2km for urban clusters, 5km for rural clusters, and 10km for an additional 1% of rural clusters to maintain participant confidentiality [23]. Shapefiles used in all maps presented were extracted from GADM.org.

Vector surfaces from the Malaria Atlas Project [24] were included in secondary analyses (**Figure S5**) as human biting rates and subsequent risk of infection are related to the zoophilic vs. anthropophilic behaviors of vectors in an area [5]. Clusters were assigned a vector species (or species combination) based on the majority type of raster cell values within a 5km buffer around each cluster. Land cover values for each survey cluster (**Figure S6**) were extracted from the European Space Agency Climate Change Initiative Land Cover Project’s “CCI LAND COVER – S2 prototype Land Cover 20m map of Africa 2016” [25]. The most common value of the raster cells which fell within a 5km buffer around each cluster were assigned as the primary land type, excluding values which represented open water and vegetation aquatic or regularly flooded areas due to sparse data.

To assess temporal variability in the association between livestock ownership and malaria infection, surveys were classified into three periods by year: 2009-2013, 2014-2018, and 2019-2022 (**Figure S7**). For surveys that collected data across multiple years, survey year was classified as the year included in the survey code.

### Statistical analysis

DHS and MIS surveys are conducted with a two-stage sampling design which first selects population clusters with probability proportional to size and then households within clusters, resulting in observations which are not independent [18]. We used hierarchical logistic regression models and survey weights to account for this nested structure and to allow for spatially varying coefficients. Models were run using a Bayesian framework with Integrated Nested Laplace Approximation (INLA) and stochastic partial differential equations (SPDE) [26,27].

Three models were run evaluating the association between livestock ownership on malaria infection: 1) a non-spatial model accounting for cluster-level random effects and survey-level random effects assumed to vary independently across clusters and surveys, 2) a spatially varying intercept model which borrowed information from neighboring clusters, and 3) a spatially varying intercept and slope model which allows for spatially varying associations between livestock ownership on malaria prevalence. Spatially structured random effects that help account for spatial confounding were modeled as Gaussian random fields with Matérn covariance functions. All models were adjusted for sex, residence, wealth, and bed net use. Model comparison was conducted using Deviance Information Criterion (DIC) metrics. All models were run using the *INLA* (*v24*.*05*.*01-1*) package in R 4.4.2 (R Foundation for Statistical Computing, Vienna, Austria). All code used for data manipulation, analysis, and models can be found at https://github.com/htopazian/animalariaSSA.

## RESULTS

### Malaria prevalence, animal ownership, and covariates

Our dataset included 69 DHS, MIS, and AIS surveys from countries within Africa which tested individuals for malaria with RDTs. After confirming that exposure, outcome, and spatial data were present, 66 surveys from 26 countries from years 2009 to 2022 were included (**Table S1**). After the removal of individuals who were not tested with RDTs, the final dataset comprised of 25,585 clusters of households and 392,843 individuals. Countries contributed a median of 2 (IQR: 1 to 4) surveys, with a median of 332 (IQR: 206 to 531) clusters and 5,300 (IQR: 4,053 to 7,268) participants. Thirty-five (53.0%) surveys were DHS, 29 (43.9%) were MIS, and two (3.0%) surveys were AIS. The national proportion of RDT+ individuals among the study population ranged from 0.004 (SE: 0.002) in The Gambia in 2019 to 0.74 (SE: 0.01) in Burkina Faso in 2010 and the proportion owning livestock ranged from 0.09 (SE: 0.01) in Gabon in 2019 to 0.86 (SE: 0.01) in Burkina Faso in 2010. The proportion of respondents living in rural areas ranged from 0.11 (SE: 0.01) in Gabon in 2019 to 0.92 (SE: 0.01) in Madagascar in 2016 and the proportion sleeping under bed nets ranged from 0.09 (SE: 0.01) in Mauritania in 2020 to 0.84 (SE: 0.01) in Niger in 2021. The majority (58, 87.9%) of surveys focused on children 6-59 months, with the remaining testing a wider age range.

The prevalence of malaria and the prevalence of animal ownership by type varied within countries and between countries, with chicken/poultry and goat ownership having a broad consistent distribution across the continent, while ownership of cattle, horses/donkeys/mules, pigs, and sheep occurred in regional pockets (**Figure 1**). Missing data across all surveys included chickens/poultry: 2,480 individuals (0.6%); goats: 830 (0.2%); horses/donkeys/mules: 31,192 (8.0%); pigs: 102,169 (26.0%); and sheep: 669 (0.2%).

**Figure 1.**
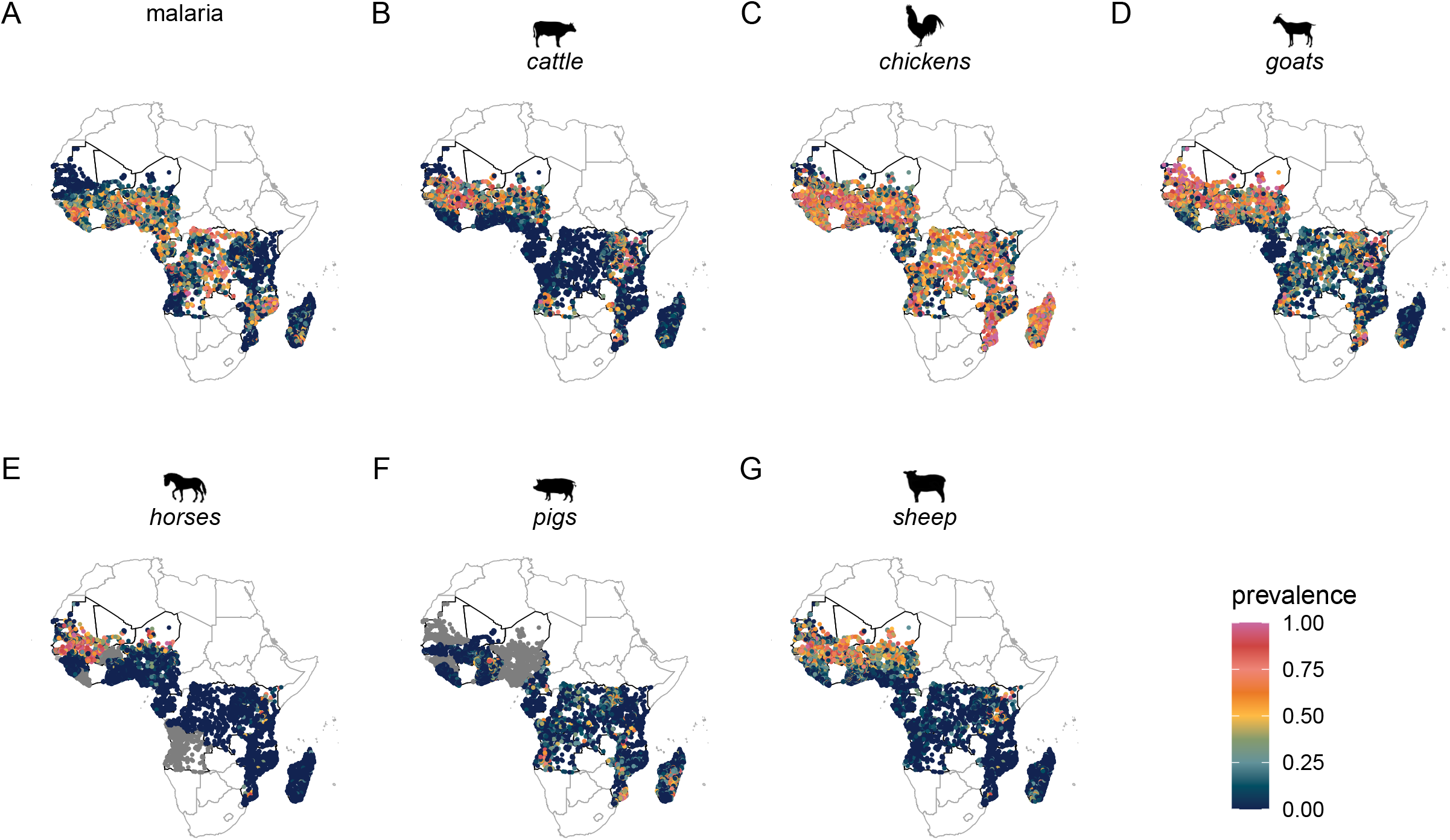
Prevalence of malaria via rapid diagnostic test (A) and prevalence of animal ownership by animal type (B-G). The most recent survey from each country is represented, comprised of 11,238 clusters from 26 Demographic and Health Surveys or Malaria Indicator Surveys from years 2013 to 2022 are represented. Gray points indicate survey clusters where information on ownership of the select animal was unavailable (horses/donkeys/mules: 1,289 (11.5%); pigs: 2,660 (23.7%)).

The median number of animals owned by individual households was low: 4 cattle (IQR: 2 to 10), 8 chickens (IQR: 4 to 15), 4 goats (IQR: 2 to 8), 2 horses/donkeys/mules (IQR: 1 to 3), 2 pigs (IQR: 1 to 3), and 4 sheep (IQR: 2 to 9) (**Figure 2**). The number of animals owned showed regional variation by animal type; larger groups of cattle were found in the Sahel region, Tanzania, Mozambique, and Angola than elsewhere on the continent (**Figure S8**). Large numbers of goats were found in the Sahel region, West Africa, Tanzania, Kenya, Mozambique, and Angola. Herds of horses/donkeys/mules were found in the Sahel region as well as in Tanzania and Northern Kenya. Small pockets of areas owned 2+ pigs in Senegal, Mozambique, Uganda, Togo, Madagascar, Benin, Burkina Faso, and Angola.

**Figure 2.**
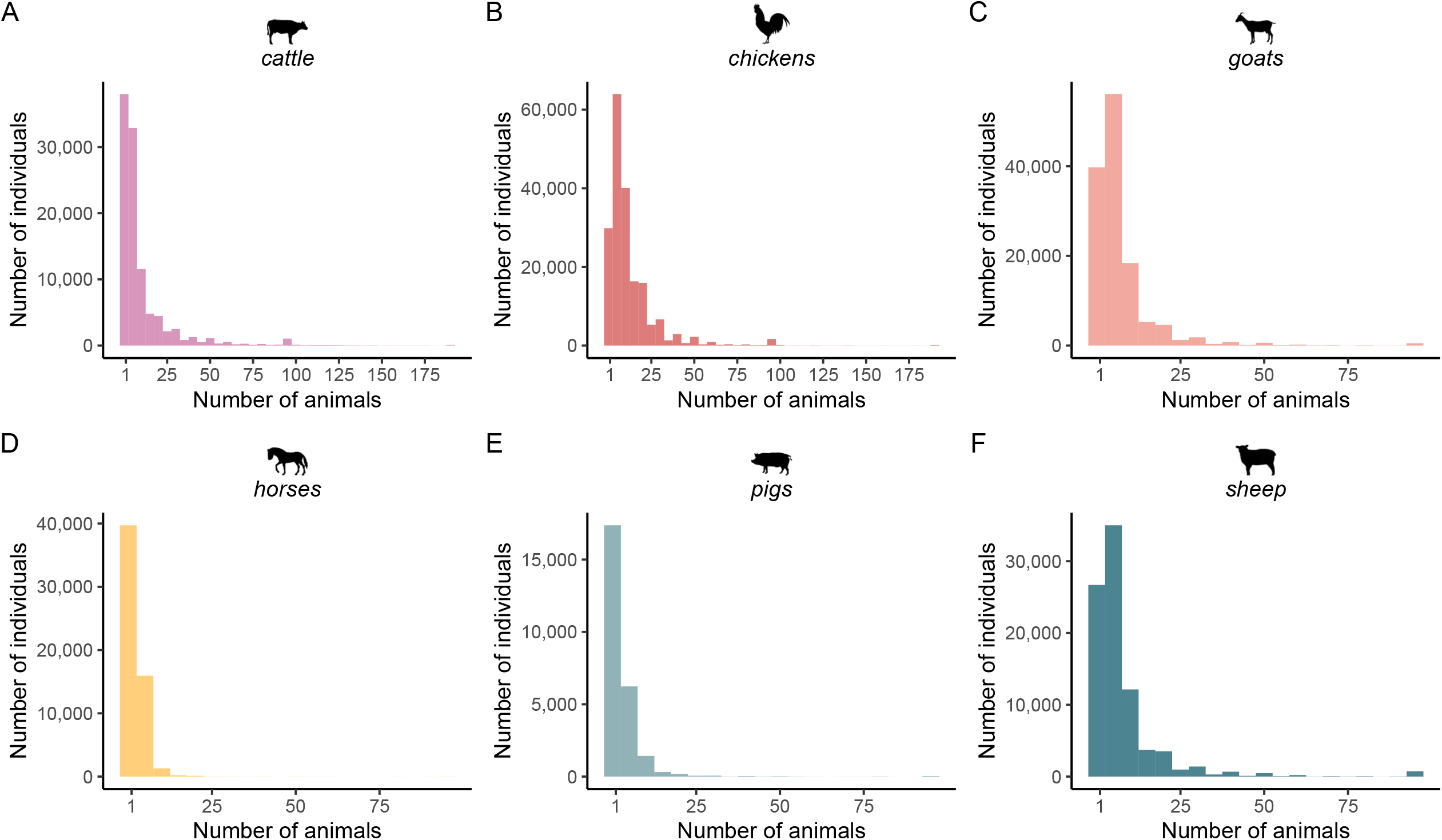
Number of animals owned, stratified by survey by animal type (A-F). 66 surveys from years 2009 to 2022 are represented.

Large flocks of sheep were owned in the Sahel region and in Benin, Kenya, and Tanzania, and chicken flock sizes were more evenly distributed across the study area. Of all survey clusters included in the analysis, 65.7% were in areas with all three dominant vector species; 11.1% were in areas with *An. funestus* and *An. gambiae*, 10.9% were in areas with *An. arabiensis* and *An. funestus*, 4.8% in areas with *An. funestus* only, 2.2% in areas with *An. arabiensis* only, 1.6% in areas with *An. gambiae* only, and 3.8% were in areas missing vector species information. Of all survey clusters, 42.1% were classified as cropland, 24.4% as tree cover areas, 13.4% as grassland, 13.5% as built-up areas, 4.7% as shrubs cover areas, and 1.9% as bare areas.

### Models

INLA models were run using three specifications: a non-spatial model, a spatially varying intercept model, and a spatially varying slope model. Adding a spatially varying intercept to the non-spatial models shifted odds ratio estimates farther from the null for each animal type (**Table 1**). Spatial intercept models showed harmful overall effects for ownership of cattle (OR: 1.67, 95% UI: 1.54, 1.80), chickens/poultry (OR: 1.67, 95% UI: 1.55, 1.81), goats (OR: 1.63, 95% UI: 1.51, 1.76), horses/donkeys/mules (OR: 1.55, 95% UI: 1.42, 1.70), pigs (OR: 1.61, 95% UI: 1.48, 1.76), and sheep (OR: 1.61, 95% UI: 1.49, 1.74) on malaria prevalence. Based on DIC values, the spatially varying slope model was the best fit for all models. Estimates for all animal types showed both protective and harmful effects on malaria infection depending on location (**Figure 3**). Odds ratios ranged from 0.33 to 2.59 for cattle (SD range: 1.14 to 1.61), 0.36 to 3.33 (SD: 1.09 to 1.67) for chickens/poultry, 0.38 to 3.00 (SD: 1.10 to 1.69) for goats, 0.40 to 2.52 (SD: 1.16 to 1.62) for horses/donkeys/mules, 0.42 to 2.09 (SD: 1.12 to 1.82) for pigs, and 0.50 to 1.98 (SD: 1.08 to 1.59) for sheep.

**Table 1.**
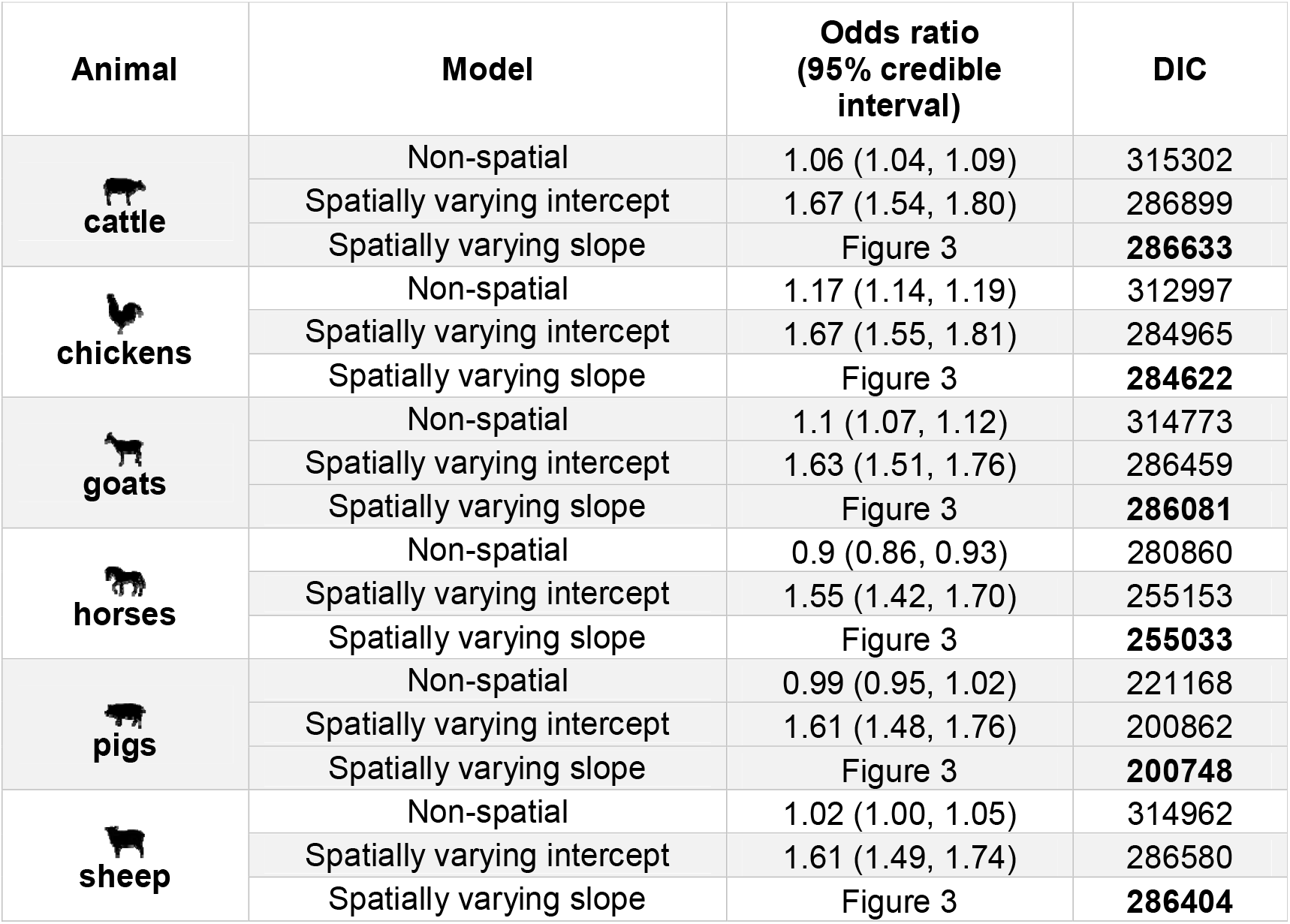
Model results for the relationship between animal ownership (yes/no) and malaria infection, stratified by animal type. Odds ratios and 95% credible intervals are presented from three models: a non-spatial model, a spatial model with a spatially varying intercept, and a spatial model with a spatially varying slope. Each model is adjusted for gender, residence (urban/rural), wealth quintile, and if an individual slept under an insecticide-treated net the night before the survey. The estimates of spatially varying slope models are not interpretable and are shown instead in **Figure 3**. Note: DIC = Deviance Information Criterion.

| Animal | Model | Odds ratio<br>(95% credible<br>interval) | DIC |
| --- | --- | --- | --- |
| cattle | Non-spatial | 1.06 (1.04, 1.09) | 315302 |
|  | Spatially varying intercept | 1.67 (1.54, 1.80) | 286899 |
|  | Spatially varying slope | Figure 3 | **286633** |
| chickens | Non-spatial | 1.17 (1.14, 1.19) | 312997 |
|  | Spatially varying intercept | 1.67 (1.55, 1.81) | 284965 |
|  | Spatially varying slope | Figure 3 | **284622** |
| goats | Non-spatial | 1.1 (1.07, 1.12) | 314773 |
|  | Spatially varying intercept | 1.63 (1.51, 1.76) | 286459 |
|  | Spatially varying slope | Figure 3 | **286081** |
| horses | Non-spatial | 0.9 (0.86, 0.93) | 280860 |
|  | Spatially varying intercept | 1.55 (1.42, 1.70) | 255153 |
|  | Spatially varying slope | Figure 3 | **255033** |
| pigs | Non-spatial | 0.99 (0.95, 1.02) | 221168 |
|  | Spatially varying intercept | 1.61 (1.48, 1.76) | 200862 |
|  | Spatially varying slope | Figure 3 | **200748** |
| sheep | Non-spatial | 1.02 (1.00, 1.05) | 314962 |
|  | Spatially varying intercept | 1.61 (1.49, 1.74) | 286580 |
|  | Spatially varying slope | Figure 3 | **286404** |

**Figure 3.**
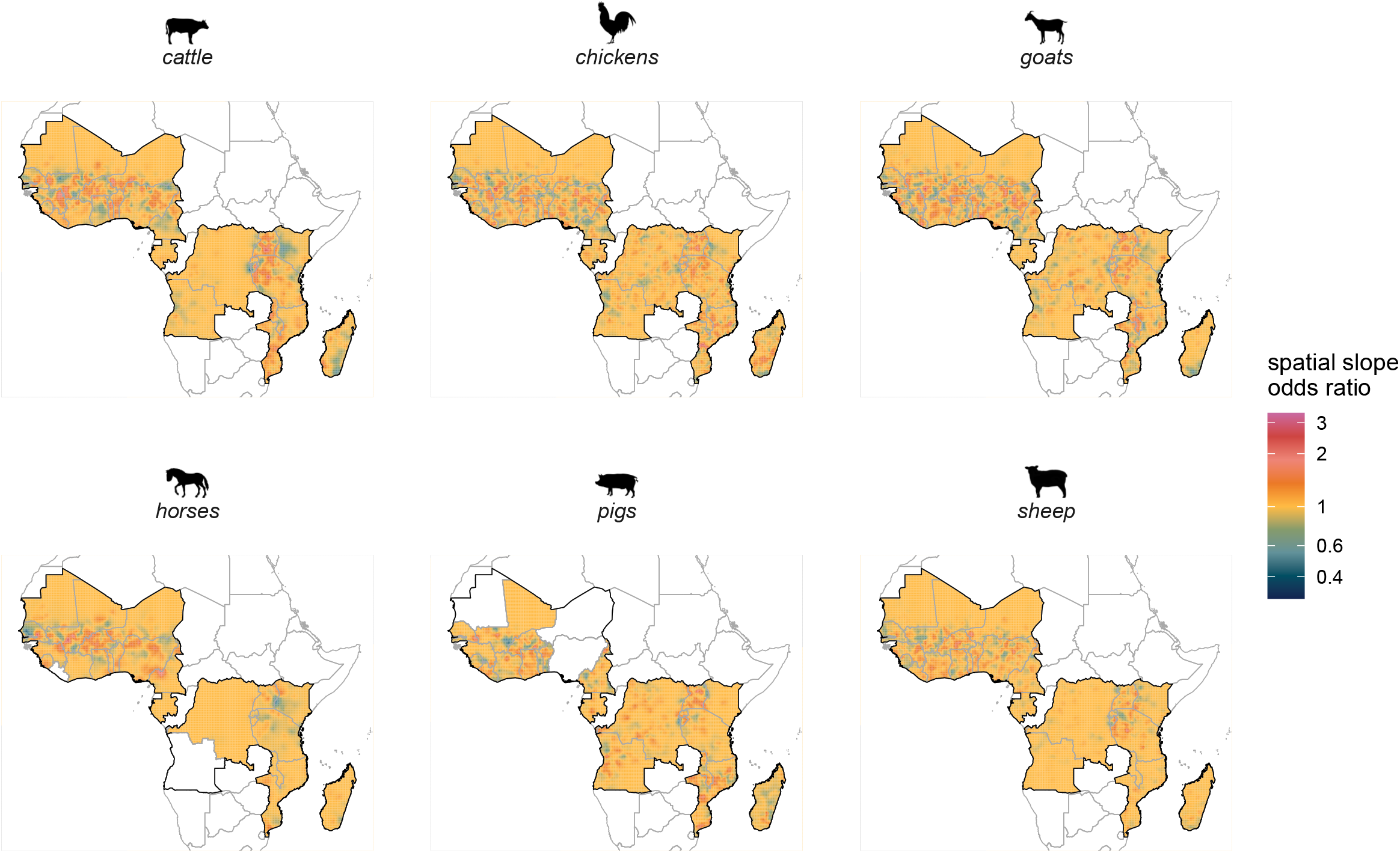
Spatially varying odds of animal ownership on malaria prevalence, stratified by animal. Standard deviation estimates are mapped in **Figure S9**.

Models stratified by time show differing patterns of spatially varying estimates of the log odds of animal ownership on malaria prevalence (**Figure S10, Figure S11**). A closer look at Senegal, the country with the most available survey timepoints, shows wider spatial variation in odds ratios from surveys conducted between 2009-2013 than those in 2014-2018 and 2019-2022 (**Figure S13, Figure S14**). The higher transmission country of Mozambique also shows changing estimates across time, even within animal group (**Figure S15, Figure S16**).

Stratifying models by the dominant vector species of each cluster using spatial intercept models showed varying associations by animal type and vector species (**Figure 4A, Table S2**). Estimates for each animal for areas with mixed species combinations were all greater than areas with a single dominant vector species, ranging from 1.13 (95% UI: 0.65, 1.95) for horses/donkeys/mules in areas with *An. funestus* and *An. gambiae*, to 2.47 (95% UI: 2.01, 3.05) for pigs in areas with *An. arabiensis* and *An. funestus*. Areas classified as *An. arabiensis* had a lower estimate for pigs (OR: 0.80, 95% UI: 0.21, 3.07. Areas classified as *An. funestus* had a lower estimate for horses/donkeys/mules (OR: 0.97, 95% UI: 0.38, 2.49), and areas classified as *An. gambiae* had lower estimates for horses (OR: 0.16, 95% UI: 0.02, 1.32) and sheep (OR: 1.05, 95% UI: 0.55, 2.00) although uncertainty intervals were wide. Stratifying models by the number of animals owned by households showed little variation across all animal types (**Figure 4B, Table S2**). Stratifying models by the type of land cover surrounding households showed the highest estimates for areas classified as bare cover across all animal types except pigs (where data were sparse), ranging from 2.54 (95% UI: 1.42, 4.56) for chickens/poultry to 4.22 (95% UI: 2.24, 7.96) for cattle, however uncertainty intervals were wide (**Figure 4C, Table S2**). Estimates were lowest in built-up areas for horses/donkeys/mules (OR: 1.16. 95% UI: 0.76, 1.78) and in areas with tree cover.

**Figure 4.**
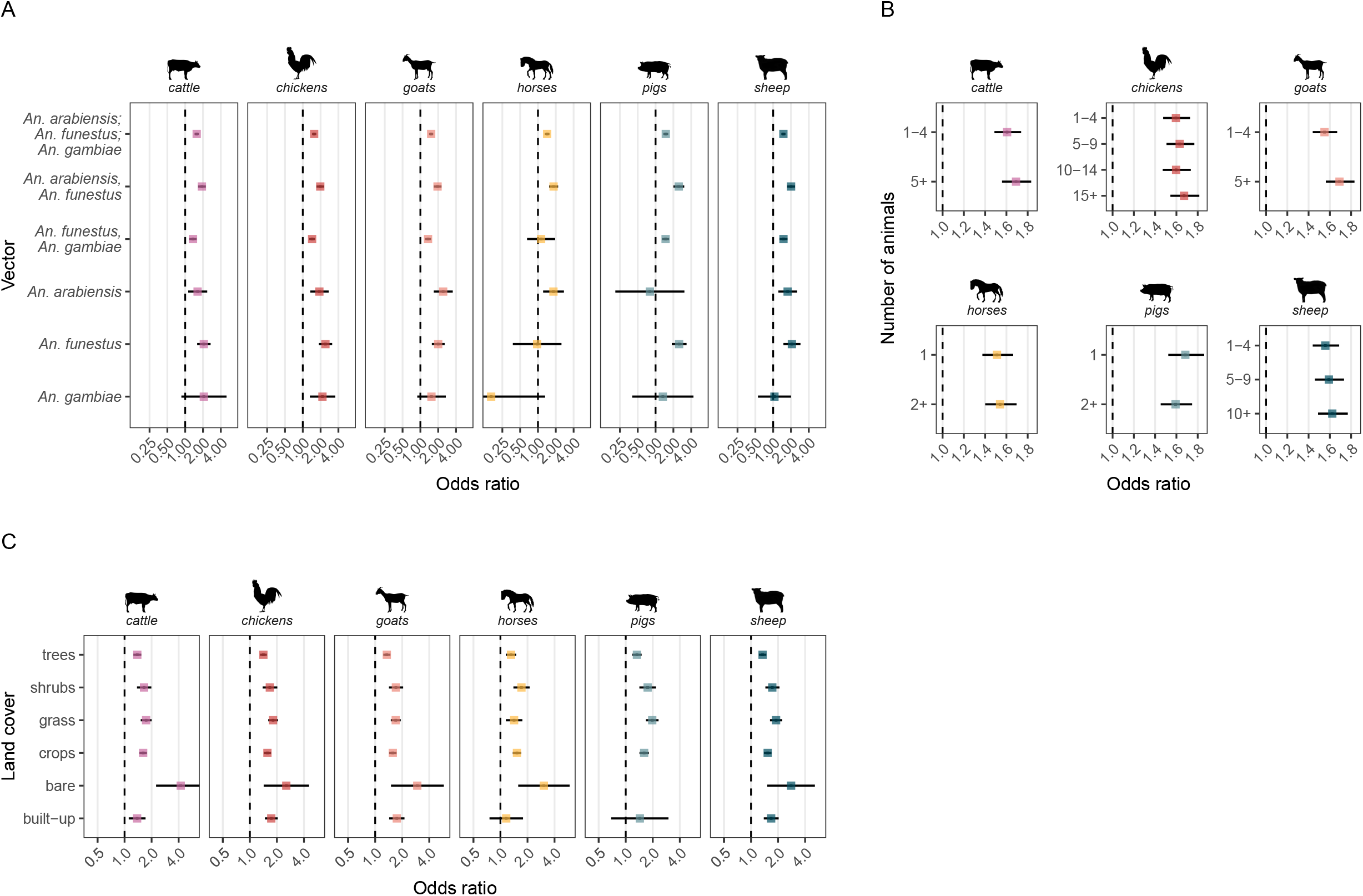
INLA model results for the relationship between animal ownership (yes/no) and malaria prevalence, stratified by animal type and A) vector species, B) number of animals owned, and C) land cover. INLA models have random intercepts for DHS survey cluster and survey ID, and a spatially varying intercept. Each model is adjusted for gender, residence (urban/rural), wealth quintile, and if an individual slept under an insecticide-treated net the night before the survey. Vector species data are extracted from an *Anopheles* distribution surface,[24] data on the number of animals owned are taken from DHS surveys, and land cover data are extracted from the European Space Agency Climate Change Initiative Land Cover project’s 2016 Africa land cover surface.[25]

## DISCUSSION

Our analysis of 392,843 individuals across 66 DHS surveys and 14 years shows spatial and temporal variation in the association between livestock ownership and malaria infection at the population level. Spatially varying slope models showed subnational variation in estimates, with both positive and negative associations even within the same country. Our results did not provide evidence that modification by vector species, number of animals owned, and land cover explain variations across the continent. Maps of spatially varying malaria and livestock associations highlight local areas where zooprophylaxis may offer benefit after accounting for these factors.

Outcomes from this study support evidence for the mixed associations between household animal ownership and malaria prevalence found in the literature. For example, in Zambia, owning cattle and other animal types was associated with a reduced risk of malaria [15], while in Pakistan, the presence of domestic cattle indoors was associated with a higher parasite prevalence [28], and in the Gambia, sleeping proximity to cattle had no effect on malaria prevalence.^15^ In addition to finding varying associations over geographical space, our analysis also shows differing patterns by animal type; the same sub-region may demonstrate protective effects from one species but harmful effects from another. This is a known phenomenon [14], hypothesized to stem from local vector biting preferences and the nature of human-animal interactions. Modelling studies provide support for both zooprophylaxis and zoopotentiation; livestock function as alternative blood meal sources which both divert mosquitoes away from humans, but also sustain mosquito densities [17].

Our analysis also shows variation in estimates stratified by vector species, animal herd/flock size, and type of land cover. Two reviews of published zooprophylaxis research concluded that ownership of domesticated animals in areas with *An. gambiae* and *An. funestus* could lead to increased malaria risk, but may be protective in areas with zoophilic *An. arabiensis* where livestock are housed far from human housing at night [10,29]. Here, we also observe lower odds ratios in areas with *An. arabiensis* as compared to areas with *An. funestus* and *An. gambiae* when examining cattle and chickens/poultry ownership. Although the literature reports general trends, we know that extrinsic environmental factors are also likely at play, such as local host availability and presence of other malaria control interventions.

In the Democratic Republic of the Congo, Zambia, and Tanzania, the protective effect of cattle ownership is modified by the number of animals owned [14,15,30]. In this analysis we see small variations in the outcome by animal type and number of animals owned across all surveys, but no substantial differences at the continent level. Modeling has shown that for malaria, increasing the number of animals has little zoorophylactic effect, but changing human accessibility to mosquito biting is influential [17]. This suggests that the number of animals owned could also be a proxy for human/animal interactive behavior, with larger or smaller herd sizes correlated with proximity to humans.

No known existing research has examined modification by land cover. In our analysis we found that areas classified as bare cover had the highest odds ratio estimates between animal ownership and malaria prevalence, perhaps due to the closer proximity of human settlements and animals in arid or desert-like settings, providing both breeding sites and blood meal sources.

Our analysis found not only geographical variation in the association between animal ownership and malaria prevalence, but also temporal variation, a trend which has not been previously examined. We hypothesize that the temporal variation seen could stem in part from changing human animal interactions across the seasons, as DHS data collection often took several months to complete, or the changing nature of human/animal interactions over years, as farming practices modernized.

This analysis has several limitations stemming from the constraints of available data. Surveys were conducted at different times of year, and human interactions with livestock also change seasonally, as has been documented in Zimbabwe, with cattle herded farther away from villages during rainy seasons to protect crops [30]. Any seasonal changes in the association between livestock ownership and malaria prevalence are not captured in this analysis of cross-sectional data. Our dataset primarily consists of children under 5 years, and we could not assess variation by age group. DHS surveys also do not collect information on proximity of animals to sleeping areas or characteristics of participant’s interactions with livestock, information which is also rarely found in the literature, but speculated to be an important factor to effective zooprophylaxis. We do not have continent-wide vector subspecies data, but variable biting preferences has been observed within broader species complexes [5], and changes in biting behavior over time could be important to assess. We did not investigate combination effects of owning more than one type of animal, but this may make teasing out animal type modifications more difficult.

Relationships between animals and humans are complex and dynamic, and change both across geographies and over time. Characterizing the nature of human interactions with animals will be essential for future studies investigating zoophrophylactic strategies. Given that some vector species feed on both humans and animals, personal protection measures for humans such as insecticide-treated bed nets and indoor residual spraying may not be enough to eliminate residual transmission. Tools must address alternative sources of blood meals, including endectocides to kill mosquitoes feeding on livestock, and veterinary insecticides [13,31]. Furthermore, rapid large-scale environmental, demographic, and socioeconomic changes across Africa may necessitate longitudinal monitoring of evolving interactions between humans, animals, and the environment over time. Our findings suggest the presence of an important and localized relationship between livestock, humans, and vectors, highlighting the need for an integrated One Health approach involving stakeholders from human, veterinary, and planetary health fields in decision-making for malaria control and prevention.

## CRediT AUTHORSHIP CONTRIBUTION STATEMENT

**Hillary M Topazian**: Conceptualization, Data curation, Formal analysis, Visualization, Writing – original draft, Writing – review and editing

**Camille E Morgan**: Conceptualization, Writing – reviewing and editing

**Varun Goel**: Conceptualization, Writing – reviewing and editing

## DATA AVAILABILITY

All relevant data are within the manuscript and supplement. Data that support study findings are available for download from the DHS MEASURE website, conditional on approval from DHS. All code to replicate data analysis and spatial models can be found at https://github.com/htopazian/animalariaSSA.

## ACKNOWLEDGEMENT

We thank the participants of the DHS surveys included in this analysis for their continued contributions to research and the DHS field teams who collected the original data.

