## Supplemental Information for "Spatial and temporal associations between animal ownership and malaria prevalence in Africa using cross-sectional national Demographic and Health Surveys"

### TECHNICAL METHODS

*INLA model structure*

We used a Bayesian framework with Integrated Nested Laplace Approximation (INLA) and stochastic partial differential equations (SPDE),[1,2] to explore the relationship between household animal ownership and malaria. Three models were run: 1) a non-spatial model accounting for cluster-level and survey-level random effects, 2) a spatially varying intercept model, and 3) a spatially varying intercept and slope model.

*Cluster-level random effects*

Our first model assessed the relationship between animal ownership and malaria after adjusting for covariates and accounting for intra-cluster correlation among survey clusters and within surveys:

$$logit(p_{ij}) = x_i\beta + u_j + v_j$$

$$u \sim N(0, \sigma_u^2 I)$$

$$v \sim N(0, \sigma_v^2 I)$$

Here, we assess the log-odds of malaria prevalence, $p_{ij}$, for individual $i$ in cluster $j$ using $x_i$ to represent a vector of covariates at the individual level, $\beta$ as the corresponding regression coefficients, $u_j$ as the cluster-level varying random intercept, and $v_j$ as the survey-level varying random intercept to control for residual confounding within correlation structures.

*General spatial confounding*

The second model included a spatially varying intercept to adjust for spatial confounding, requiring approximation of a continuous spatial field using a stochastic partial differential equation (SPDE)-based approach.[2] To create the spatial field, we used a mesh of discrete spatial points informed by survey cluster locations (**Figure S1** shows the overall mesh, **Figure S2** shows meshes by time period). Distinct mesh surfaces were developed for each model and tested at varying levels of granularity to balance resolution and computational constraints. The spatially varying intercept model was defined as:

$$logit(p_{ij}) = x_i\beta + u_j + v_j + s_j$$

$$u \sim N(0, \sigma_u^2 I)$$

$$v \sim N(0, \sigma_v^2 I)$$

$$s \sim N(0, \sigma_s^2 \Sigma(\phi))$$

Spatial random intercepts, $s$, are defined by priors which borrow from neighboring survey sites with an exponential decay over increasing distance using a SPDE model with a Matérn covariance structure to capture spatial autocorrelation. Priors had a median spatial range of 1.8 degrees (~200 km at the equator) with a 50% probability that the range parameter was greater than 1.8, and a median standard deviation of $\sqrt{0.2}$ with a 10% probability that the standard deviation was greater than $\sqrt{0.2}$. These values were chosen after examining variograms for each survey to assess the extent of spatial dependence present in the data (**Figure S3**). The model also included random intercepts for survey cluster and survey.

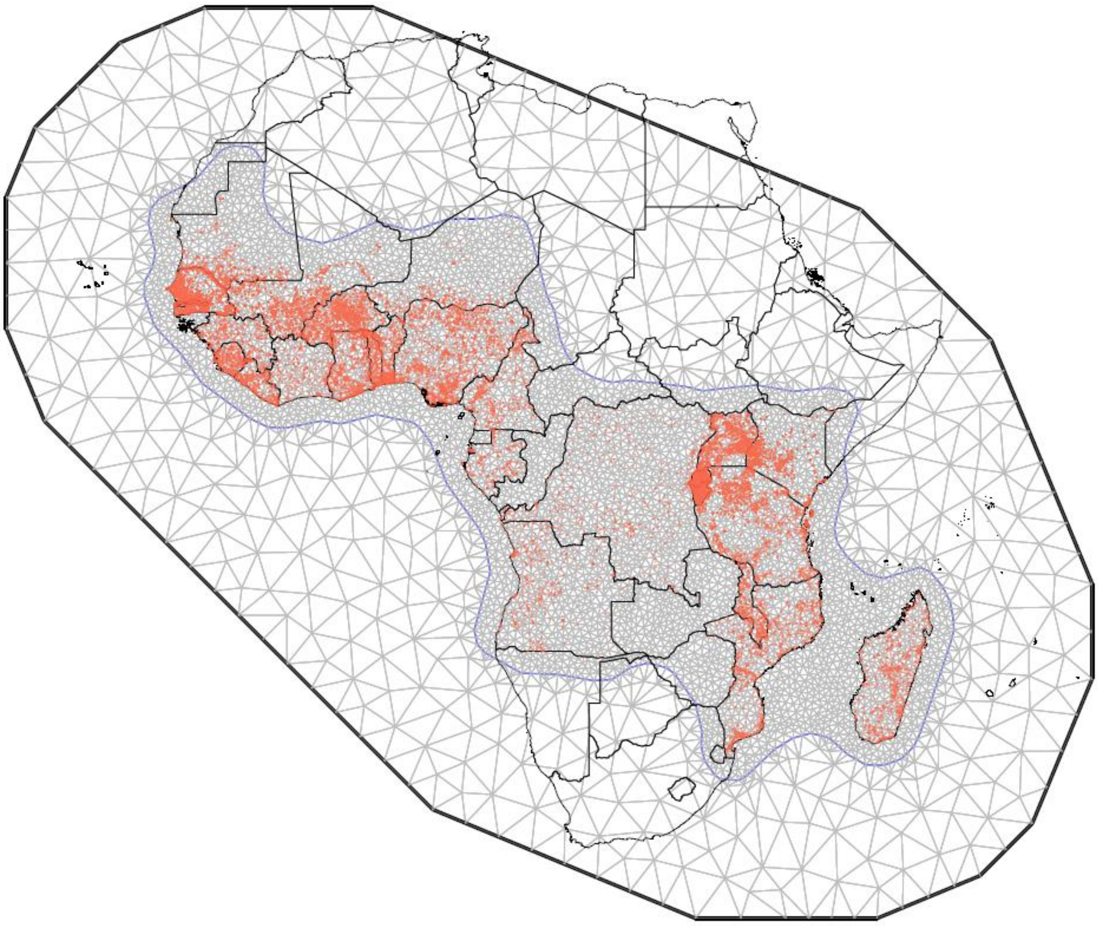

**Figure S1**. Spatial mesh formations for all DHS clusters with data used in the analysis. Clusters are marked as red points.

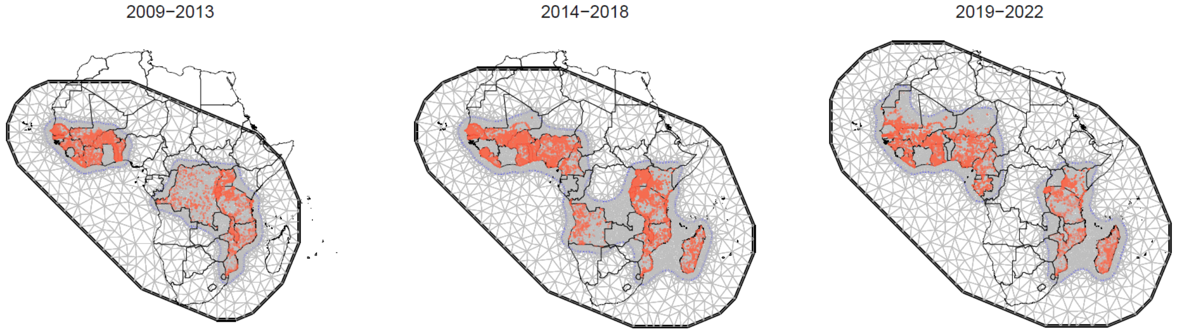


**Figure S2**. Spatial mesh formations for DHS clusters with data used in the analysis, stratified by time period. Clusters are marked as red points.

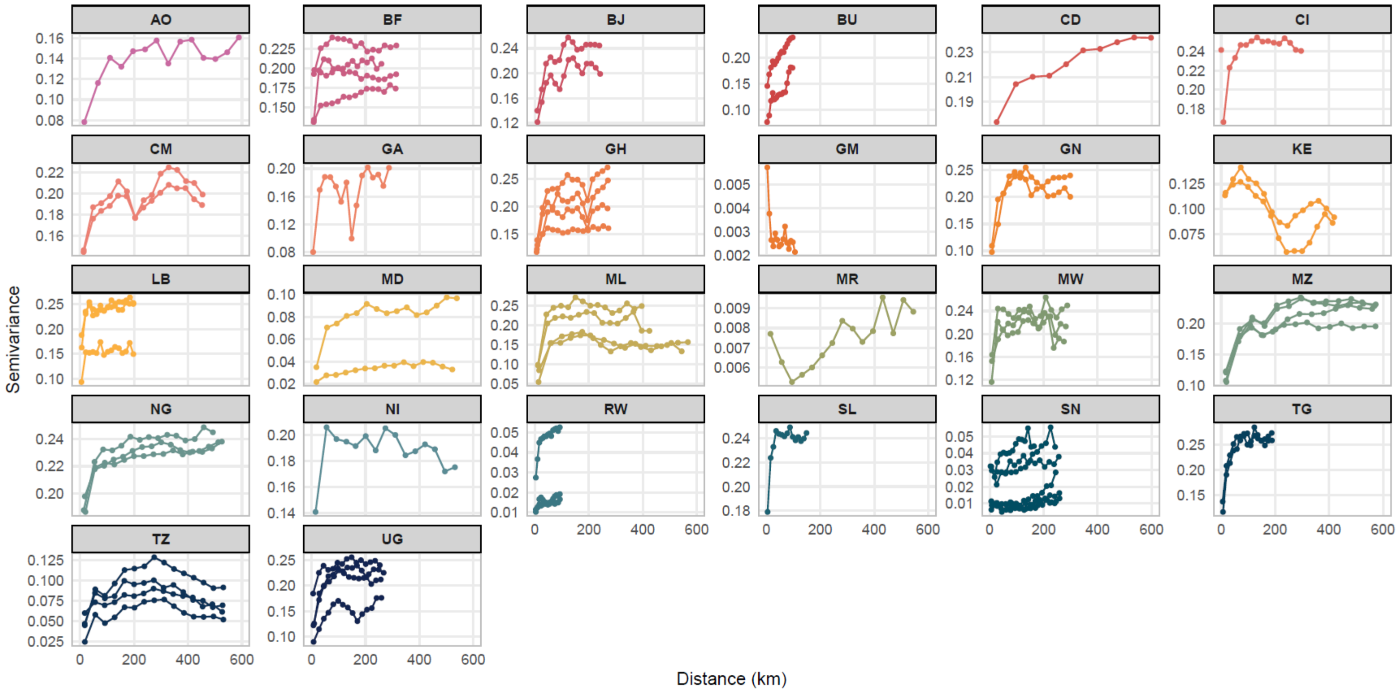


**Figure S3**. Variograms showing the spatial dependence present in survey datasets for the outcome variable of malaria infection (defined as rapid diagnostic test +) as a function of distance. Plots are stratified by country, with lines representing individual surveys by year.

*Spatial variability in the exposure-outcome relationship*

The third model additionally included spatially varying slopes to flexibly model variation in the relationship between animal ownership and malaria across space:

$$logit(p_{ij}) = x_i\beta + u_j + v_j + s_j + z_j\eta_j$$

$$u \sim N(0, \sigma_u^2 I)$$

$$v \sim N(0, \sigma_v^2 I)$$

$$s \sim N(0, \sigma_s^2 \Sigma(\phi))$$

$$\eta \sim N(0, \sigma_\eta^2 \Sigma(\phi_\eta))$$

Where $z_j$ is the cluster-level covariate of interest (animal ownership) and $\eta_j$ is the spatially varying random effect for all individuals in the specified cluster. The survey cluster and survey random intercepts and spatial intercept remain as in previous models to control for intra-cluster and intra-survey correlation and spatial confounding.

*Temporal variability in the exposure-outcome relationship*

Surveys were conducted across multiple years, spanning from the Uganda 2009 MIS to the Mozambique 2022-2023 DHS (**Table S1**). To assess temporal variability in the association between livestock ownership and malaria infection, surveys were classified into three periods by year: 2009-2013, 2014-2018, and 2019-2022 (**Figure S7**). Surveys in which data were collected over multiple years spanning these times periods were classified by the year contained in their survey code.

*Survey selection, variable classification, and missing data*

Country names were labeled as they were defined in the DHS program, with The Gambia referred to as "Gambia" and the Democratic Republic of the Congo referred to as "Congo Democratic Republic." The DHS standard survey template generally includes counts for the number of cattle, cows/bulls, chickens/poultry, goats, horses/donkeys/mules, and sheep owned by a household. Other optional variables were country-specific and included household ownership of camels, ducks/geese/turkeys, exotic/crossbreed/local/modern cattle, grasscutters, guinea fowl, guinea pigs, pigs, rabbits, and zebus. Cattle were combined with cows/bulls and exotic/cross-breed/local/modern cattle were also included within this grouping.

Our initial dataset included 69 DHS, MIS, and AIS surveys from countries within Africa which tested individuals for malaria with RDTs. After removal of 1 survey which did not collect data on livestock ownership (MD2011MIS) and 2 surveys which could not be linked to geocoordinates (GM2013DHS, RW2017MIS), 66 surveys from 26 countries from years 2009 to 2022 were included.

**ADDITIONAL TABLES AND FIGURES**

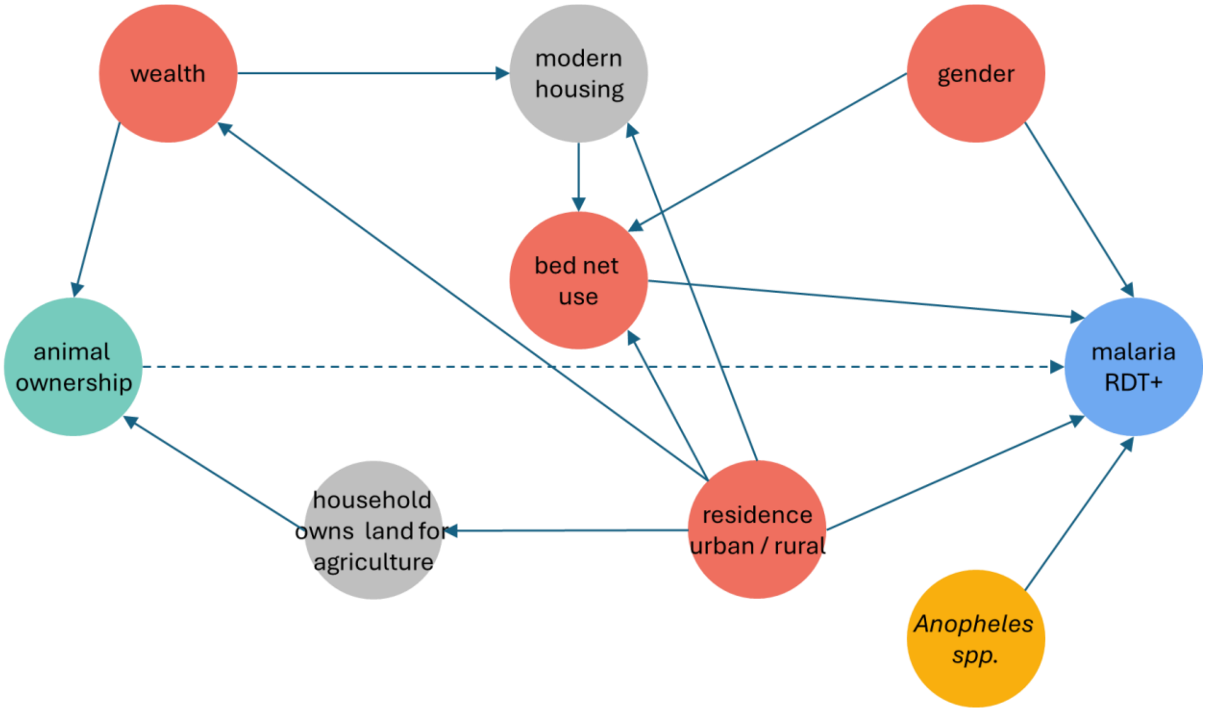


**Figure S4**. Directed Acyclic Graph showing the variables associated with the relationship between the exposure (animal ownership) and outcome (malaria). Variables are colored by type: green represents the exposure, blue represents the outcome, gray represents observed unadjusted variables, red represents variables in the adjustment set, and yellow represents variables which influence malaria prevalence but not animal ownership. The direction of the arrows indicates the direction of influence, solid arrows represent known relationships, and the dashed arrow represents the causal question of interest. The sufficient adjustment set used in our models included gender, bed net use, residence, and wealth. Note: RDT = rapid diagnostic test

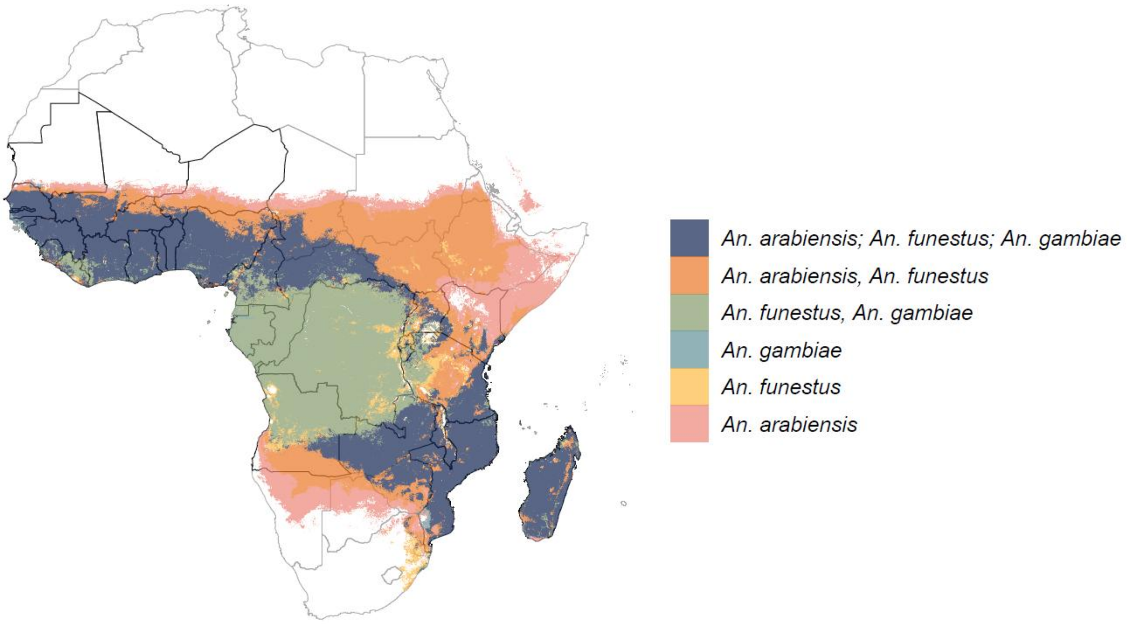


**Figure S5**. "Dominant malaria vector species globally, 2010" vector surface from the Malaria Atlas Project,[3] showing the *Anopheles* mosquito species (or species combinations) that are the most important for malaria transmission in each area.

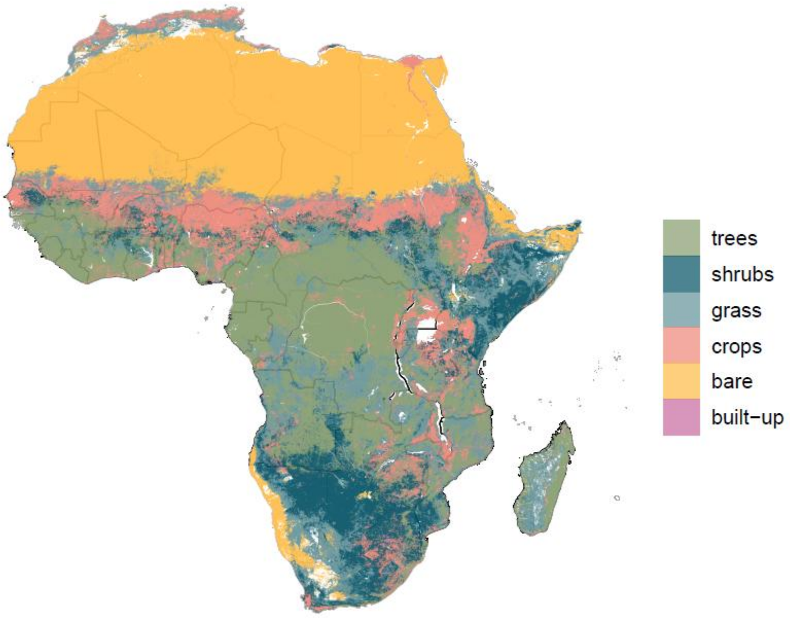


**Figure S6**. The European Space Agency Climate Change Initiative Land Cover Project's "CCI LAND COVER – S2 prototype Land Cover 20m map of Africa 2016."[4] Values which represent open water and vegetation aquatic or regularly flooded areas are not shown and were not assigned to survey cluster locations due to sparse data.

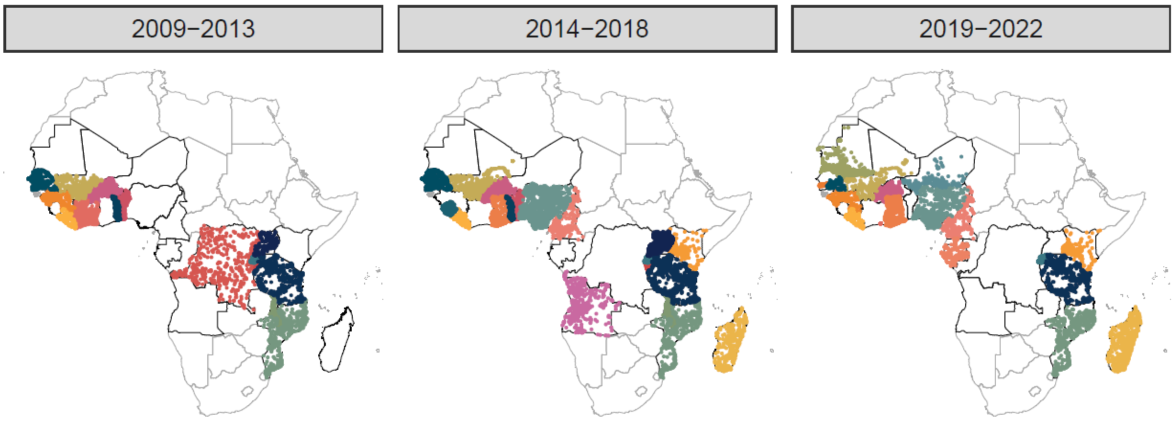


**Figure S7**. Survey cluster locations, stratified by year of survey.

**Table S1**. Characteristics of the Demographic and Health Survey data used in the analysis. Only participants with rapid diagnostic test results were included. The proportion of individuals who had a positive rapid diagnostic test and the proportion of individuals living in households owning livestock are adjusted for survey weights. Notes: AIS = Aids Indicator Survey, DHS = Demographic and Health Survey, MIS = Malaria Indicator Survey, RDT = rapid diagnostic test, SE = standard error

| Country | Survey code | Country code | Survey type | Survey year(s) | # clusters | # participants | Proportion RDT+ (SE) | Proportion living in households owning livestock (SE) | Proportion living in rural areas (SE) | Proportion who slept under an insecticide-treated net the night before the survey (SE) | Age range (years) |
|---|---|---|---|---|---|---|---|---|---|---|---|
| **Angola** | AO2015DHS | AO | DHS | 2015-2016 | 625 | 8,317 | 0.14 (0.01) | 0.33 (0.01) | 0.41 (0.01) | 0.19 (0.01) | 0 - 5 |
| **Benin** | BJ2012DHS | BJ | DHS | 2011-2012 | 741 | 4,887 | 0.25 (0.01) | 0.38 (0.01) | 0.62 (0.01) | 0.68 (0.01) | 0 - 5 |
| | BJ2017DHS | BJ | DHS | 2017-2018 | 540 | 6,118 | 0.36 (0.01) | 0.63 (0.01) | 0.62 (0.01) | 0.70 (0.01) | 0 - 5 |
| **Burkina Faso** | BF2010DHS | BF | DHS | 2010 | 541 | 5,919 | 0.74 (0.01) | 0.86 (0.01) | 0.83 (0.01) | 0.46 (0.01) | 0 - 5 |
| | BF2014MIS | BF | MIS | 2014 | 248 | 6,044 | 0.61 (0.01) | 0.77 (0.01) | 0.77 (0.02) | 0.75 (0.01) | 0 - 5 |
| | BF2017MIS | BF | MIS | 2017-2018 | 224 | 5,068 | 0.20 (0.01) | 0.83 (0.01) | 0.83 (0.01) | 0.53 (0.01) | 0 - 5 |
| | BF2021DHS | BF | DHS | 2021 | 514 | 5,767 | 0.28 (0.01) | 0.74 (0.01) | 0.76 (0.01) | 0.61 (0.01) | 0 - 5 |
| **Burundi** | BU2012MIS | BU | MIS | 2012 | 200 | 3,750 | 0.22 (0.02) | 0.66 (0.02) | 0.92 (0.02) | 0.53 (0.02) | 0 - 5 |
| | BU2016DHS | BU | DHS | 2016-2017 | 552 | 6,877 | 0.39 (0.01) | 0.67 (0.01) | 0.91 (0.01) | 0.37 (0.01) | 0 - 5 |
| **Cameroon** | CM2018DHS | CM | DHS | 2018 | 428 | 4,733 | 0.24 (0.01) | 0.50 (0.02) | 0.55 (0.02) | 0.41 (0.01) | 0 - 5 |
| | CM2022MIS | CM | MIS | 2022 | 439 | 4,295 | 0.26 (0.02) | 0.51 (0.02) | 0.57 (0.03) | 0.50 (0.02) | 0 - 5 |
| **Congo Democratic Republic** | CD2013DHS | CD | DHS | 2013-2014 | 491 | 7,533 | 0.31 (0.02) | 0.49 (0.02) | 0.68 (0.02) | 0.54 (0.02) | 0 - 5 |
| **Cote d'Ivoire** | CI2012DHS | CI | DHS | 2011-2012 | 341 | 4,581 | 0.40 (0.02) | 0.34 (0.02) | 0.63 (0.02) | 0.37 (0.02) | 0 - 46 |
| | CI2021DHS | CI | DHS | 2021 | 537 | 4,931 | 0.37 (0.01) | 0.37 (0.01) | 0.52 (0.01) | 0.44 (0.01) | 0 - 5 |
| **Gabon** | GA2019DHS | GA | DHS | 2019-2021 | 390 | 5,982 | 0.13 (0.01) | 0.09 (0.01) | 0.11 (0.01) | 0.16 (0.01) | 0 - 5 |
| **Gambia** | GM2019DHS | GM | DHS | 2019-2020 | 279 | 3,668 | 0.004 (0.002) | 0.73 (0.02) | 0.35 (0.02) | 0.44 (0.02) | 0 - 5 |
| **Ghana** | GH2014DHS | GH | DHS | 2014 | 419 | 3,151 | 0.38 (0.02) | 0.47 (0.02) | 0.54 (0.02) | 0.41 (0.02) | 0 - 5 |
| | GH2016MIS | GH | MIS | 2016 | 192 | 2,910 | 0.28 (0.02) | 0.45 (0.02) | 0.56 (0.03) | 0.51 (0.02) | 0 - 5 |
| | GH2019MIS | GH | MIS | 2019 | 192 | 2,731 | 0.23 (0.02) | 0.50 (0.02) | 0.58 (0.02) | 0.53 (0.02) | 0 - 5 |
| | GH2022DHS | GH | DHS | 2022 | 615 | 4,481 | 0.16 (0.01) | 0.47 (0.01) | 0.51 (0.01) | 0.50 (0.01) | 0 – 5 |

| Country | Survey code | Country code | Survey type | Survey year(s) | # clusters | # participants | Proportion RDT+ (SE) | Proportion living in households owning livestock (SE) | Proportion living in rural areas (SE) | Proportion who slept under an insecticide-treated net the night before the survey (SE) | Age range (years) |
|---|---|---|---|---|---|---|---|---|---|---|---|
| **Guinea** | GN2012DHS | GN | DHS | 2012 | 300 | 3,215 | 0.47 (0.02) | 0.58 (0.02) | 0.75 (0.01) | 0.26 (0.01) | 0 - 5 |
| | GN2021MIS | GN | MIS | 2021 | 169 | 4,029 | 0.33 (0.02) | 0.59 (0.02) | 0.74 (0.02) | 0.35 (0.02) | 0 - 5 |
| **Kenya** | KE2015MIS | KE | MIS | 2015 | 245 | 10,072 | 0.13 (0.01) | 0.74 (0.02) | 0.72 (0.02) | 0.48 (0.01) | 0 - 14 |
| | KE2020MIS | KE | MIS | 2020 | 297 | 11,587 | 0.07 (0.01) | 0.69 (0.02) | 0.72 (0.02) | 0.34 (0.02) | 0 - 14 |
| **Liberia** | LB2011MIS | LB | MIS | 2011 | 150 | 3,187 | 0.45 (0.02) | 0.37 (0.02) | 0.61 (0.04) | 0.35 (0.02) | 0 - 5 |
| | LB2016MIS | LB | MIS | 2016 | 150 | 2,788 | 0.45 (0.02) | 0.48 (0.02) | 0.48 (0.03) | 0.43 (0.02) | 0 - 5 |
| | LB2022MIS | LB | MIS | 2022 | 150 | 2,927 | 0.17 (0.01) | 0.48 (0.03) | 0.51 (0.03) | 0.48 (0.02) | 0 - 5 |
| **Madagascar** | MD2016MIS | MD | MIS | 2016 | 358 | 6,931 | 0.05 (0.01) | 0.67 (0.01) | 0.92 (0.01) | 0.72 (0.01) | 0 - 5 |
| | MD2021DHS | MD | DHS | 2021 | 647 | 5,874 | 0.08 (0.01) | 0.71 (0.01) | 0.85 (0.01) | 0.36 (0.01) | 0 - 5 |
| **Malawi** | MW2012MIS | MW | MIS | 2012 | 140 | 2,115 | 0.43 (0.03) | 0.56 (0.02) | 0.87 (0.01) | 0.54 (0.03) | 0 - 5 |
| | MW2014MIS | MW | MIS | 2014 | 140 | 1,921 | 0.37 (0.04) | 0.59 (0.03) | 0.86 (0.02) | 0.66 (0.02) | 0 - 5 |
| | MW2017MIS | MW | MIS | 2017 | 148 | 2,479 | 0.37 (0.02) | 0.47 (0.02) | 0.87 (0.01) | 0.27 (0.02) | 0 - 5 |
| **Mali** | ML2012DHS | ML | DHS | 2012-2013 | 413 | 5,830 | 0.47 (0.01) | 0.69 (0.01) | 0.82 (0.01) | 0.66 (0.01) | 0 - 5 |
| | ML2015MIS | ML | MIS | 2015 | 177 | 7,302 | 0.32 (0.02) | 0.77 (0.02) | 0.81 (0.01) | 0.70 (0.01) | 0 - 5 |
| | ML2018DHS | ML | DHS | 2018 | 328 | 4,125 | 0.19 (0.01) | 0.67 (0.01) | 0.80 (0.01) | 0.73 (0.01) | 0 - 5 |
| | ML2021MIS | ML | MIS | 2021 | 216 | 8,635 | 0.19 (0.01) | 0.79 (0.01) | 0.80 (0.01) | 0.65 (0.01) | 0 - 5 |
| **Mauritania** | MR2020DHS | MR | DHS | 2019-2021 | 1,181 | 10,128 | 0.01 (0.002) | 0.65 (0.01) | 0.62 (0.01) | 0.09 (0.01) | 0 - 10 |
| **Mozambique** | MZ2011DHS | MZ | DHS | 2011 | 603 | 4,913 | 0.38 (0.02) | 0.60 (0.01) | 0.72 (0.01) | 0.35 (0.01) | 0 - 5 |
| | MZ2015AIS | MZ | AIS | 2015 | 304 | 4,476 | 0.40 (0.02) | 0.53 (0.02) | 0.75 (0.01) | 0.45 (0.02) | 0 - 5 |
| | MZ2018MIS | MZ | MIS | 2018 | 221 | 4,347 | 0.39 (0.02) | 0.48 (0.02) | 0.73 (0.02) | 0.72 (0.02) | 0 - 5 |
| | MZ2022DHS | MZ | DHS | 2022-2023 | 605 | 4,185 | 0.31 (0.02) | 0.45 (0.01) | 0.73 (0.01) | 0.38 (0.01) | 0 - 5 |
| **Niger** | NI2021MIS | NI | MIS | 2021 | 206 | 4,724 | 0.29 (0.02) | 0.77 (0.01) | 0.84 (0.01) | 0.84 (0.01) | 0 - 5 |
| **Nigeria** | NG2015MIS | NG | MIS | 2015 | 322 | 5,902 | 0.44 (0.02) | 0.57 (0.01) | 0.65 (0.01) | 0.43 (0.01) | 0 - 5 |
| | NG2018DHS | NG | DHS | 2018 | 1,374 | 11,139 | 0.36 (0.01) | 0.51 (0.01) | 0.56 (0.01) | 0.43 (0.01) | 0 - 5 |
| | NG2021MIS | NG | MIS | 2021 | 567 | 10,848 | 0.39 (0.01) | 0.56 (0.01) | 0.73 (0.01) | 0.38 (0.01) | 0 - 5 |

| Country | Survey code | Country code | Survey type | Survey year(s) | # clusters | # participants | Proportion RDT+ (SE) | Proportion living in households owning livestock (SE) | Proportion living in rural areas (SE) | Proportion who slept under an insecticide-treated net the night before the survey (SE) | Age range (years) |
|---|---|---|---|---|---|---|---|---|---|---|---|
| **Rwanda** | RW2010DHS | RW | DHS | 2010 | 492 | 11,880 | 0.02 (0.002) | 0.60 (0.01) | 0.86 (0.01) | 0.63 (0.01) | 0 - 49 |
| | RW2015DHS | RW | DHS | 2014-2015 | 492 | 10,891 | 0.06 (0.004) | 0.55 (0.01) | 0.82 (0.01) | 0.66 (0.01) | 0 - 49 |
| | RW2019DHS | RW | DHS | 2019-2020 | 500 | 11,049 | 0.02 (0.002) | 0.49 (0.01) | 0.81 (0.02) | 0.44 (0.01) | 0 - 49 |
| **Senegal** | SN2010DHS | SN | DHS | 2010-2011 | 383 | 4,627 | 0.03 (0.004) | 0.70 (0.02) | 0.62 (0.04) | 0.29 (0.02) | 0 - 5 |
| | SN2012DHS | SN | DHS | 2012-2013 | 200 | 7,316 | 0.04 (0.005) | 0.75 (0.02) | 0.67 (0.02) | 0.41 (0.02) | 0 - 5 |
| | SN2014DHS | SN | DHS | 2014 | 197 | 6,646 | 0.01 (0.003) | 0.76 (0.02) | 0.58 (0.03) | 0.40 (0.02) | 0 - 5 |
| | SN2015DHS | SN | DHS | 2015 | 214 | 6,895 | 0.01 (0.001) | 0.72 (0.02) | 0.65 (0.01) | 0.55 (0.02) | 0 - 5 |
| | SN2016DHS | SN | DHS | 2016 | 214 | 6,703 | 0.01 (0.001) | 0.76 (0.02) | 0.63 (0.01) | 0.66 (0.02) | 0 - 5 |
| | SN2017DHS | SN | DHS | 2017 | 400 | 10,897 | 0.01 (0.002) | 0.77 (0.01) | 0.64 (0.01) | 0.61 (0.01) | 0 - 5 |
| | SN2020MIS | SN | MIS | 2020-2021 | 124 | 3,657 | 0.05 (0.01) | 0.85 (0.01) | 0.80 (0.02) | 0.38 (0.02) | 0 - 5 |
| **Sierra Leone** | SL2016MIS | SL | MIS | 2016 | 336 | 7,666 | 0.54 (0.01) | 0.56 (0.01) | 0.62 (0.02) | 0.43 (0.01) | 0 - 5 |
| **Tanzania** | TZ2012AIS | TZ | AIS | 2011-2012 | 573 | 7,516 | 0.09 (0.01) | 0.75 (0.01) | 0.84 (0.01) | 0.69 (0.01) | 0 - 5 |
| | TZ2015DHS | TZ | DHS | 2015-2016 | 607 | 10,935 | 0.15 (0.01) | 0.67 (0.01) | 0.76 (0.01) | 0.50 (0.01) | 0 - 5 |
| | TZ2017MIS | TZ | MIS | 2017 | 436 | 7,164 | 0.07 (0.01) | 0.65 (0.02) | 0.73 (0.03) | 0.54 (0.01) | 0 - 5 |
| | TZ2022DHS | TZ | DHS | 2022 | 623 | 5,184 | 0.08 (0.01) | 0.57 (0.02) | 0.74 (0.02) | 0.58 (0.01) | 0 - 5 |
| **Togo** | TG2013DHS | TG | DHS | 2013 | 329 | 3,868 | 0.40 (0.02) | 0.61 (0.02) | 0.67 (0.01) | 0.41 (0.01) | 0 - 5 |
| | TG2017MIS | TG | MIS | 2017 | 171 | 3,202 | 0.44 (0.02) | 0.62 (0.02) | 0.67 (0.02) | 0.71 (0.02) | 0 - 5 |
| **Uganda** | UG2009MIS | UG | MIS | 2009 | 170 | 3,998 | 0.52 (0.03) | 0.74 (0.02) | 0.88 (0.01) | 0.66 (0.03) | 0 - 4 |
| | UG2014MIS | UG | MIS | 2014-2015 | 208 | 4,854 | 0.30 (0.02) | 0.70 (0.02) | 0.85 (0.02) | 0.74 (0.01) | 0 - 5 |
| | UG2016DHS | UG | DHS | 2016 | 681 | 5,415 | 0.31 (0.01) | 0.71 (0.01) | 0.81 (0.02) | 0.60 (0.01) | 0 - 5 |
| | UG2018MIS | UG | MIS | 2018-2019 | 316 | 7,058 | 0.17 (0.01) | 0.64 (0.02) | 0.77 (0.03) | 0.60 (0.01) | 0 - 5 |

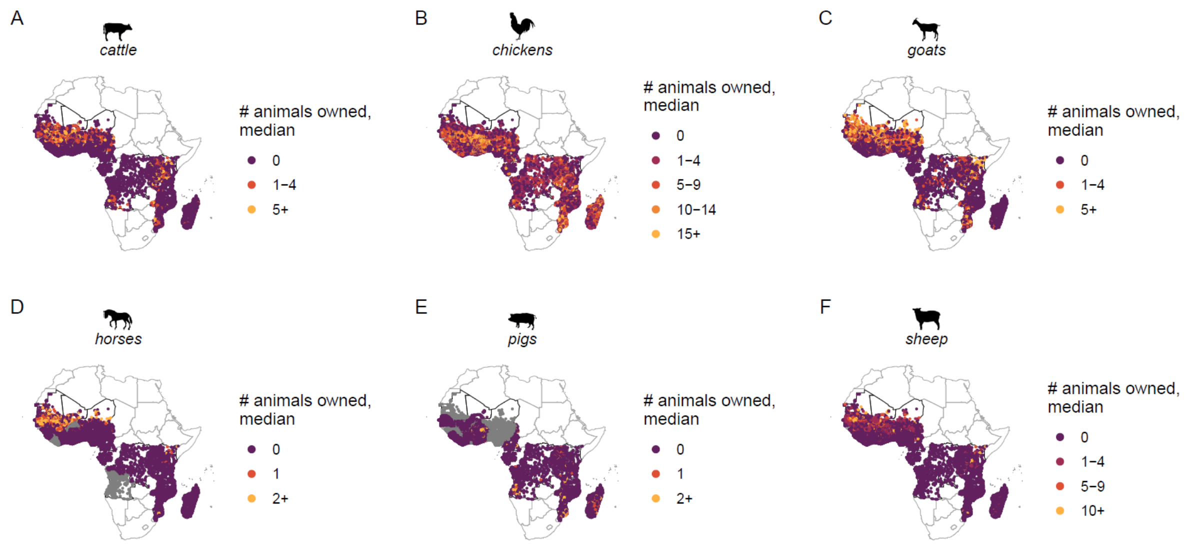


**Figure S8**. Prevalence of animal ownership by animal type and stratified by the median number of animals owned per individual (A-F). 25,585 clusters from 64 Demographic and Health Surveys or Malaria Indicator Surveys from years 2009 to 2022 are represented. Gray points indicate survey clusters where information on ownership of a particular animal was unavailable.

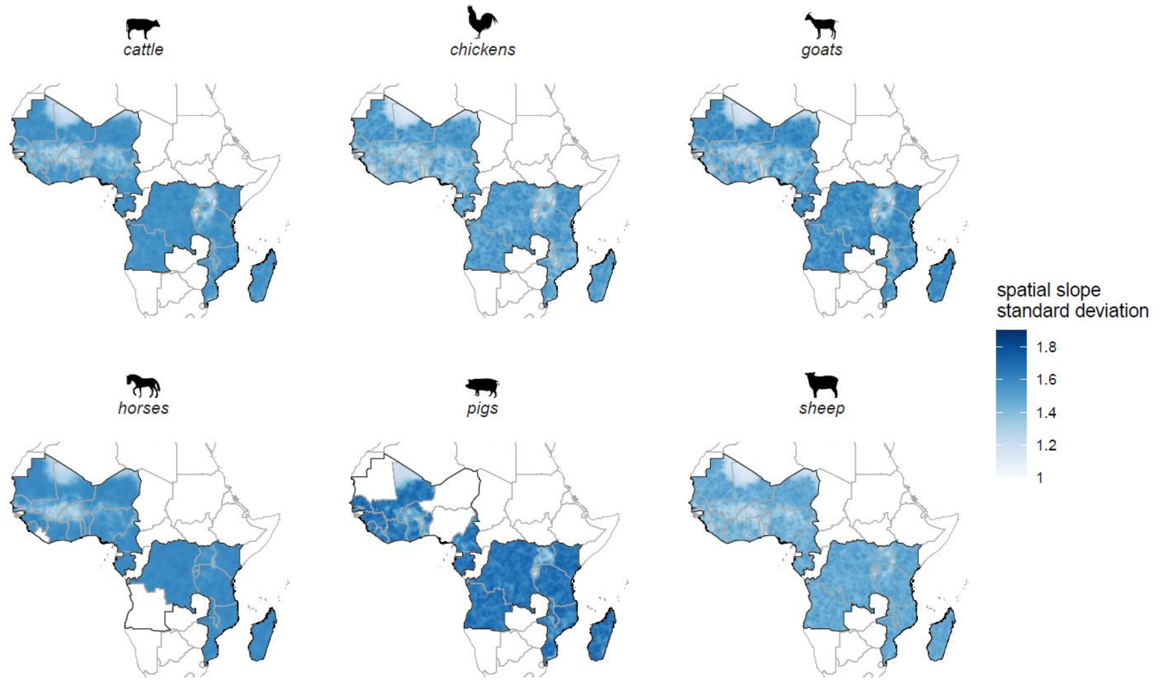


**Figure S9**. Spatially varying standard deviation of the odds of animal ownership on malaria prevalence, stratified by animal type (A-F).

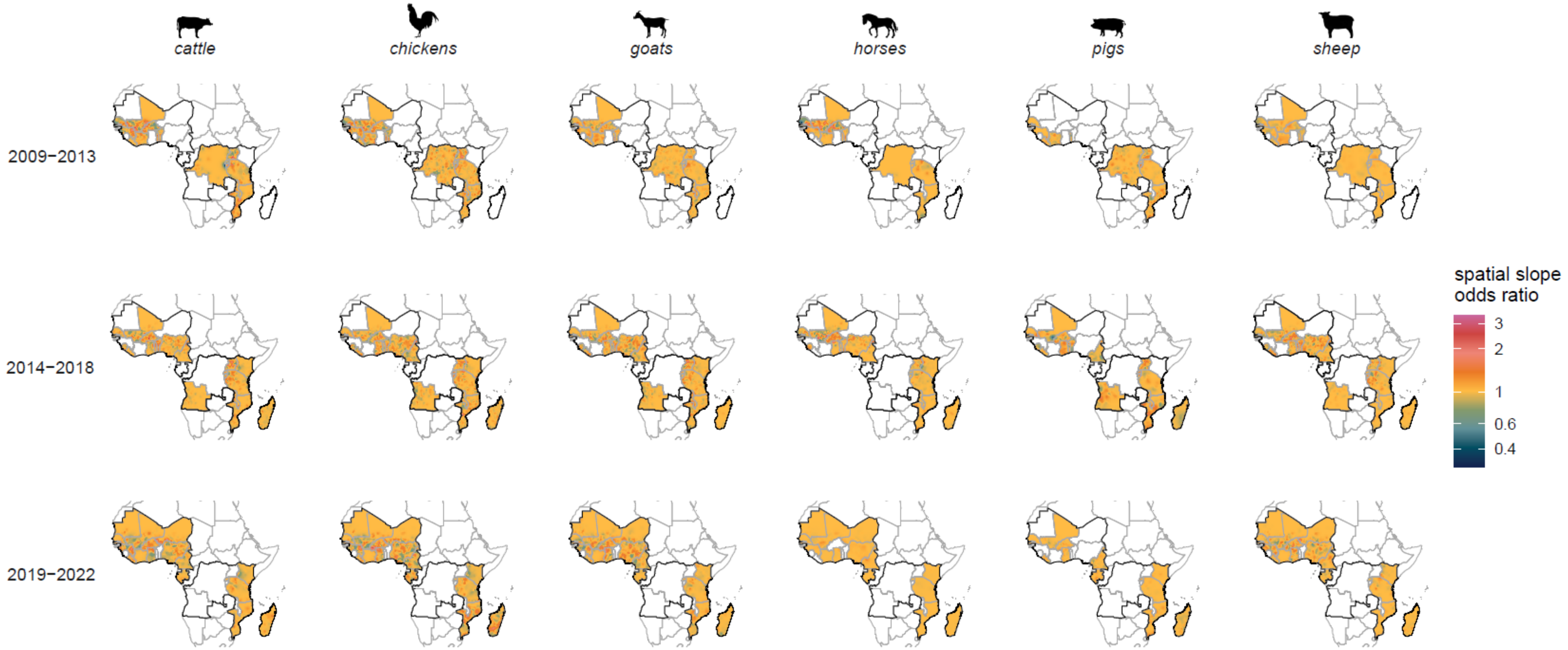


**Figure S10**. Spatially varying estimates of the odds of animal ownership on malaria prevalence, stratified by animal. Standard deviation estimates are shown in **Figure S11**.

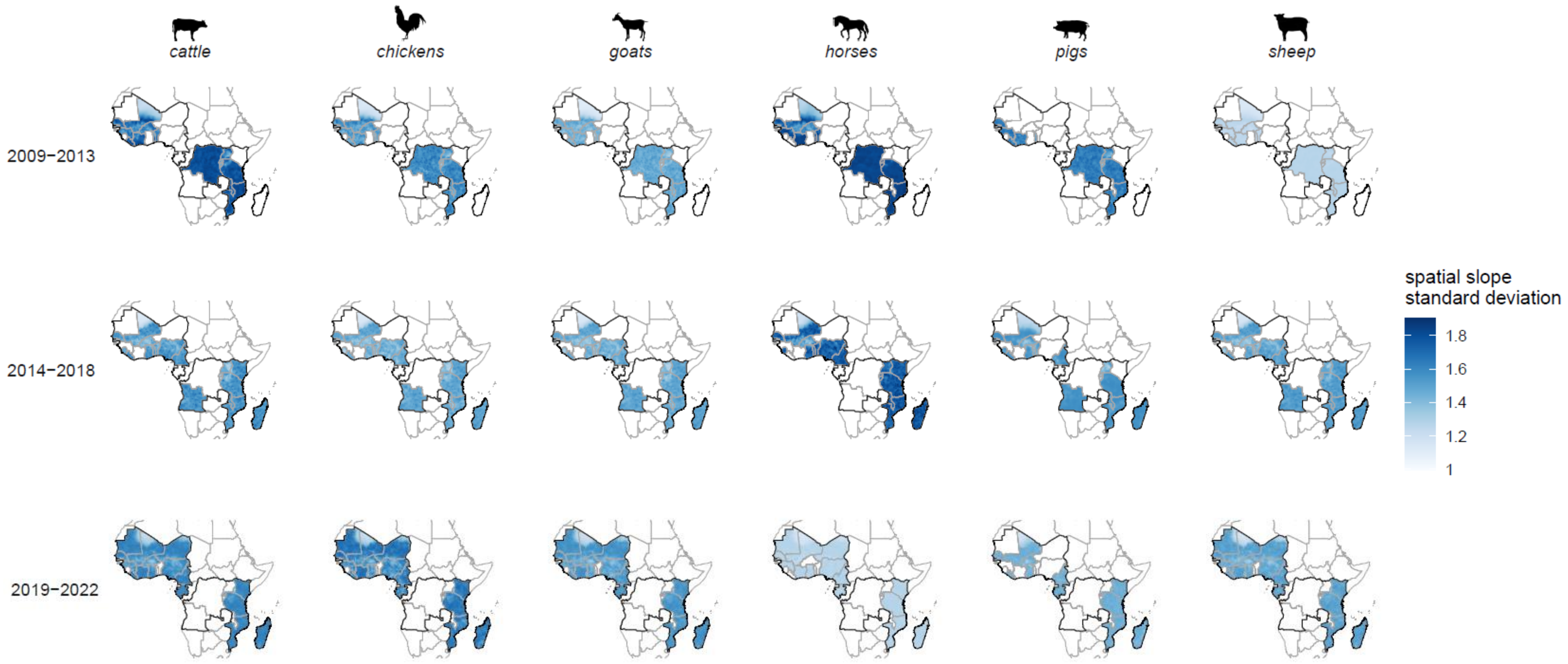


**Figure S11**. Spatially varying standard deviation of the odds of animal ownership on malaria prevalence, stratified by animal.

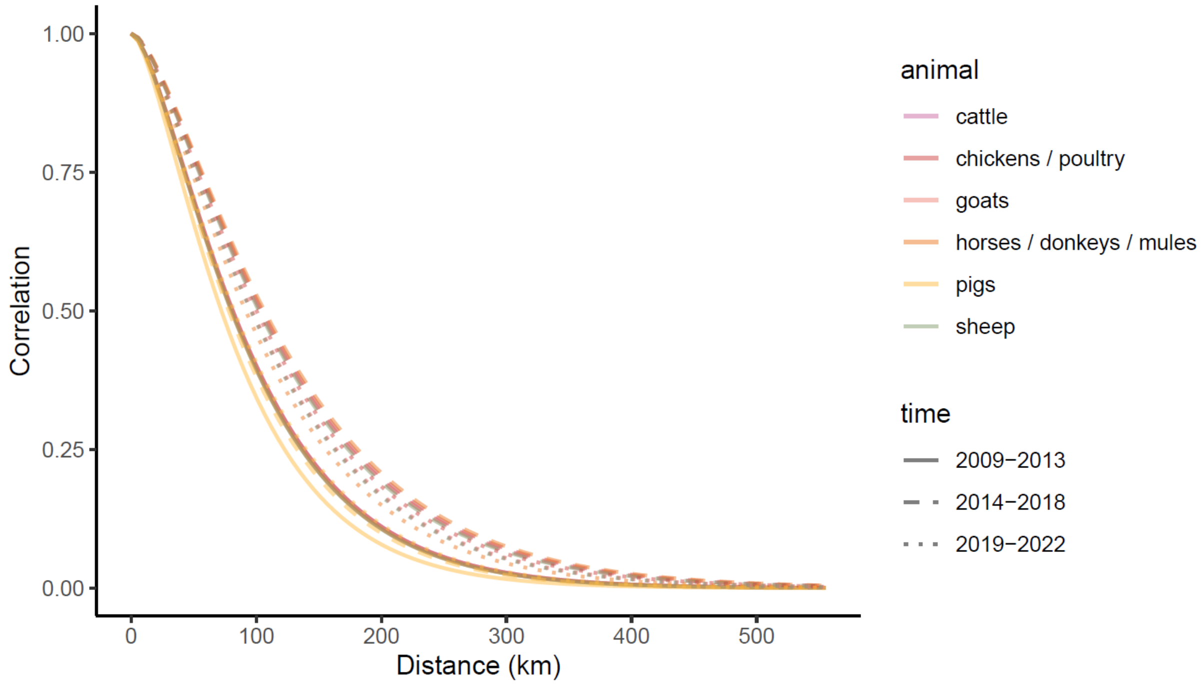


**Figure S12**. Modeled spatial correlograms using a Matérn covariance function, stratified by animal type and time.

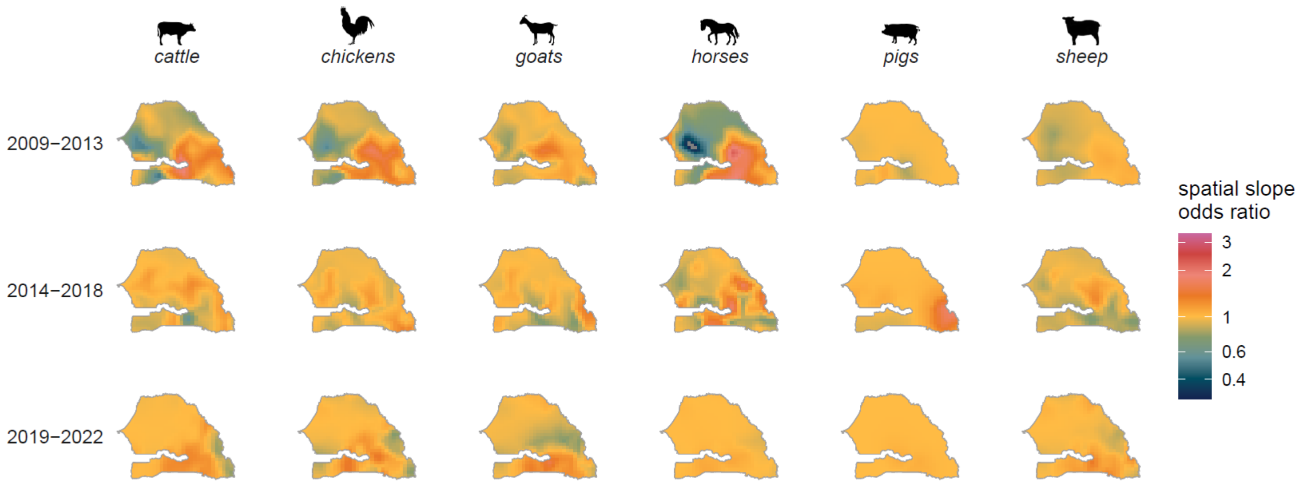


**Figure S13**. Spatially varying estimates of the odds of animal ownership on malaria prevalence in Senegal, stratified by animal type and time period. Standard deviation estimates are shown in **Figure S14**.

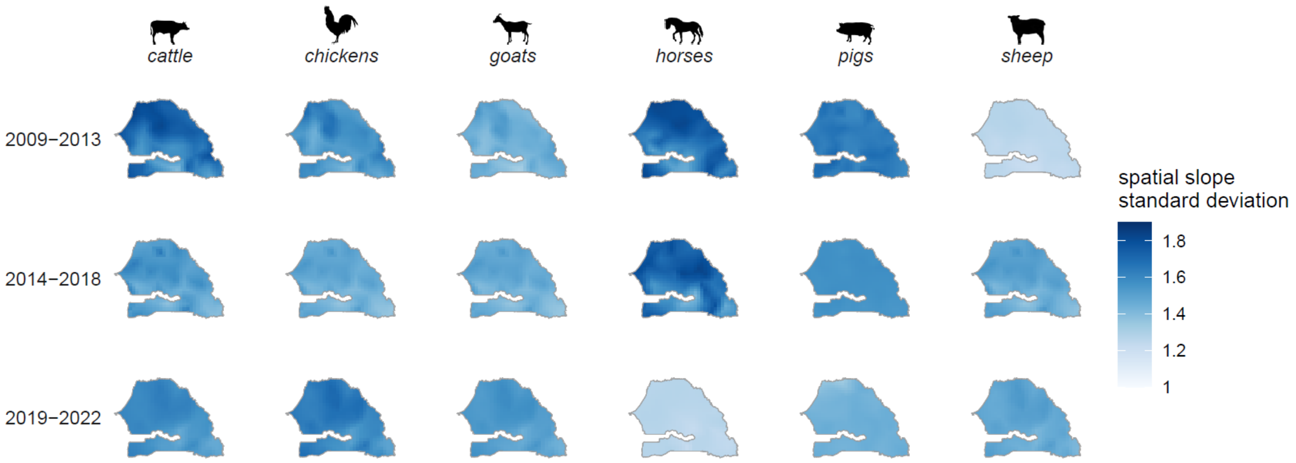


**Figure S14**. Spatially varying standard deviation of the odds of animal ownership on malaria prevalence in Senegal, stratified by animal type and time period.

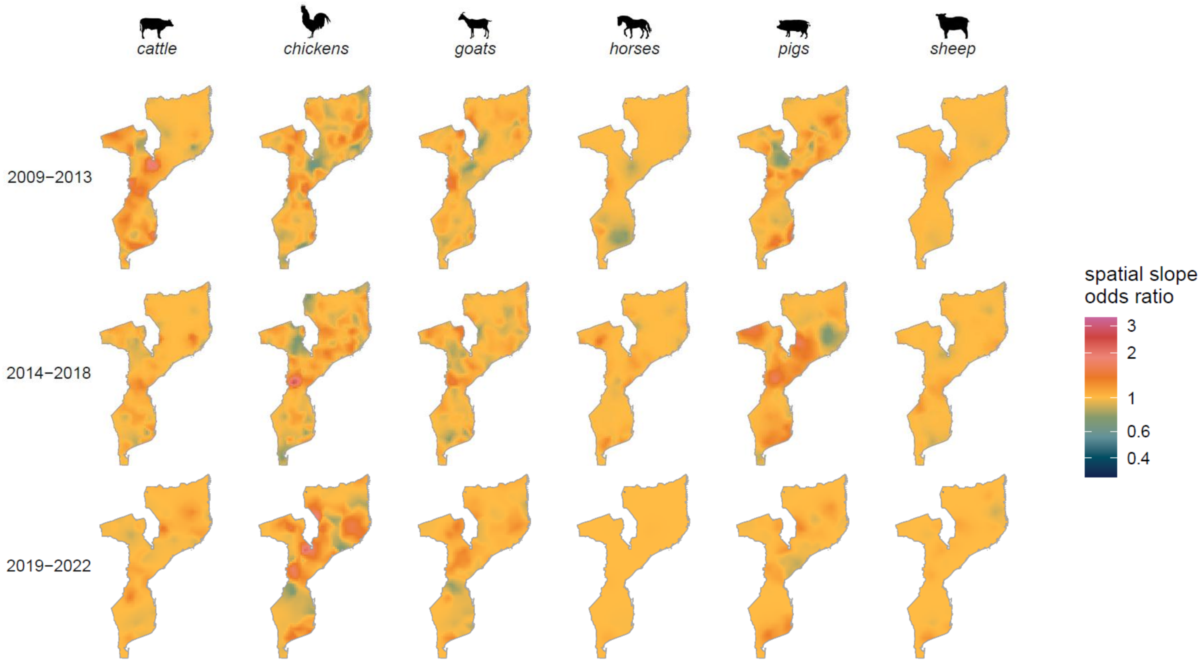


**Figure S15**. Spatially varying estimates of the odds of animal ownership on malaria prevalence in Mozambique, stratified by animal type and time period. Standard deviation estimates are shown in **Figure S16**.

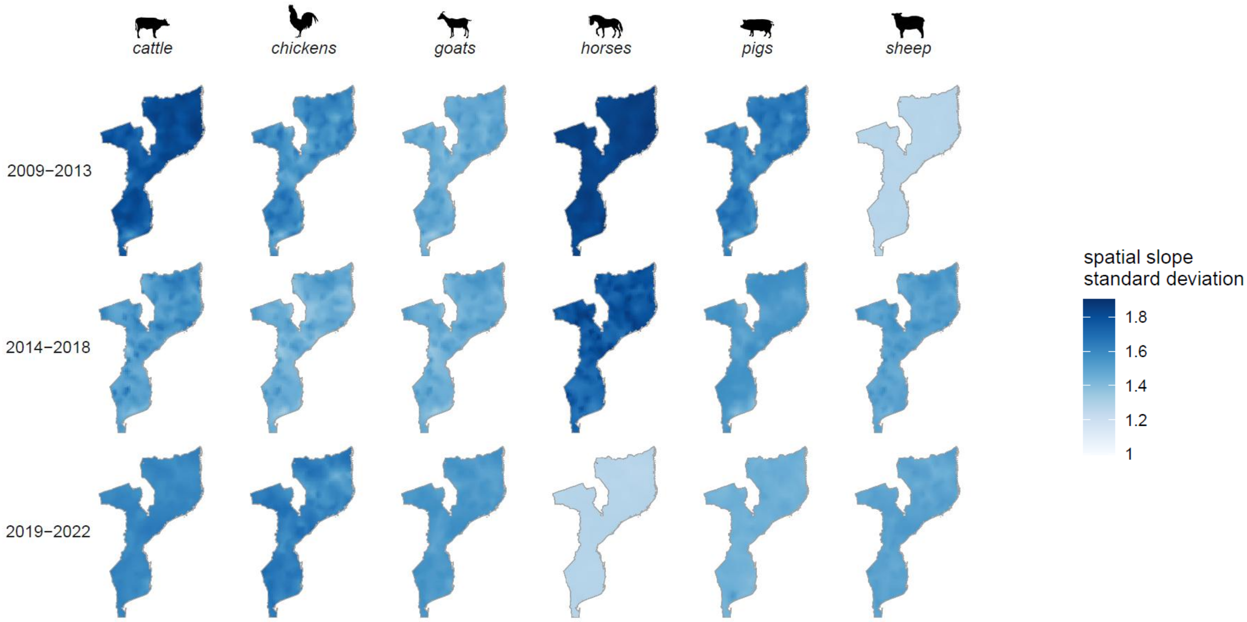


**Figure S16**. Spatially varying standard deviation of the odds of animal ownership on malaria prevalence in Mozambique, stratified by animal type and time period.

**Table S2**. Model results for the relationship between animal ownership (yes/no) and malaria infection, stratified by animal type. Estimates represent odds ratios and 95% credible intervals of spatial INLA models with a spatially varying intercept. Each model is adjusted for gender, residence (urban/rural), wealth quintile, and if an individual slept under an insecticide-treated net the night before the survey.

| Vector | Cattle | Chickens / poultry | Goats | Horses / donkeys / mules | Pigs | Sheep |
|---|---|---|---|---|---|---|
| ***An. arabiensis; An. funestus; An. gambiae*** | 1.57 (1.44, 1.70) | 1.56 (1.44, 1.69) | 1.52 (1.40, 1.65) | 1.43 (1.31, 1.57) | 1.49 (1.36, 1.64) | 1.49 (1.37, 1.62) |
| ***An. arabiensis, An. funestus*** | 1.92 (1.65, 2.23) | 1.97 (1.70, 2.3) | 1.95 (1.69, 2.27) | 1.83 (1.54, 2.19) | 2.47 (2.01, 3.05) | 2.01 (1.73, 2.34) |
| ***An. funestus, An. gambiae*** | 1.35 (1.17, 1.56) | 1.44 (1.29, 1.61) | 1.35 (1.20, 1.51) | 1.13 (0.65, 1.95) | 1.48 (1.29, 1.70) | 1.49 (1.28, 1.74) |
| ***An. arabiensis*** | 1.62 (1.13, 2.36) | 1.91 (1.34, 2.73) | 2.43 (1.69, 3.52) | 1.83 (1.22, 2.75) | 0.80 (0.21, 3.07) | 1.76 (1.22, 2.54) |
| ***An. funestus*** | 2.06 (1.60, 2.69) | 2.41 (1.87, 3.14) | 2 .00 (1.56, 2.57) | 0.97 (0.38, 2.49) | 2.50 (1.88, 3.34) | 2.07 (1.49, 2.89) |
| ***An. gambiae*** | 2.07 (0.86, 5.00) | 2.15 (1.32, 3.54) | 1.54 (0.89, 2.69) | 0.16 (0.02, 1.32) | 1.33 (0.40, 4.37) | 1.05 (0.55, 2.00) |

| Number of animals owned | Cattle | Chickens / poultry | Goats | Horses / donkeys / mules | Pigs | Sheep |
|---|---|---|---|---|---|---|
| **1** | - | - | - | 1.51 (1.37, 1.66) | 1.68 (1.52, 1.86) | - |
| **2+** | - | - | - | 1.54 (1.40, 1.7) | 1.59 (1.45, 1.75) | - |
| **1-4** | 1.61 (1.49, 1.74) | 1.60 (1.48, 1.73) | 1.55 (1.44, 1.67) | - | - | 1.56 (1.44, 1.69) |
| **5+** | 1.69 (1.56, 1.83) | - | 1.69 (1.56, 1.83) | - | - | - |
| **5-9** | - | 1.63 (1.51, 1.77) | - | - | - | 1.59 (1.46, 1.73) |
| **10+** | - | - | - | - | - | 1.62 (1.49, 1.77) |
| **10-14** | - | 1.60 (1.47, 1.73) | - | - | - | - |
| **15+** | - | 1.67 (1.54, 1.81) | - | - | - | - |

| Land cover | Cattle | Chickens / poultry | Goats | Horses / donkeys / mules | Pigs | Sheep |
|---|---|---|---|---|---|---|
| **trees** | 1.39 (1.25, 1.53) | 1.41 (1.30, 1.54) | 1.35 (1.23, 1.48) | 1.31 (1.15, 1.50) | 1.33 (1.18, 1.50) | 1.34 (1.22, 1.48) |
| **shrubs** | 1.65 (1.38, 1.99) | 1.67 (1.39, 2.02) | 1.70 (1.42, 2.05) | 1.71 (1.40, 2.11) | 1.75 (1.42, 2.17) | 1.72 (1.44, 2.07) |
| **grass** | 1.74 (1.51, 2.00) | 1.81 (1.60, 2.05) | 1.69 (1.49, 1.93) | 1.42 (1.15, 1.75) | 1.97 (1.68, 2.32) | 1.90 (1.63, 2.23) |
| **crops** | 1.61 (1.46, 1.77) | 1.57 (1.43, 1.72) | 1.57 (1.43, 1.73) | 1.53 (1.37, 1.70) | 1.60 (1.42, 1.81) | 1.53 (1.39, 1.69) |
| **bare** | 4.22 (2.24, 7.96) | 2.54 (1.42, 4.56) | 2.94 (1.50, 5.81) | 3.04 (1.58, 5.89) | - | 2.79 (1.52, 5.14) |
| **built-up** | 1.38 (1.11, 1.71) | 1.73 (1.47, 2.04) | 1.74 (1.43, 2.12) | 1.16 (0.76, 1.78) | 1.43 (0.69, 2.99) | 1.68 (1.39, 2.03) |
